# Hospital environments harbor chlorhexidine tolerant bacteria potentially linked to chlorhexidine persistence in the environment

**DOI:** 10.1101/2024.10.07.24315058

**Authors:** Jiaxian Shen, Yuhan Weng, Tyler Shimada, Meghana Karan, Andrew Watson, Rachel L. Medernach, Vincent B. Young, Mary K. Hayden, Erica M. Hartmann

## Abstract

A One Health approach is essential in combating antibiotic and antimicrobial resistance. Chlorhexidine (di)gluconate (CHG), a widely used antiseptic in medical intensive care units (MICU), has recently come under scrutiny. However, studies on CHG tolerance, particularly in interconnected indoor environments, are limited. We comprehensively explored CHG tolerance in MICU environments from chemical, microbial, and molecular perspectives. Using microcosm experiments and field surveys, we demonstrated that CHG, if transferred from patient skin to environments, can persist on surfaces despite cleaning and disinfection and decrease to sublethal levels for clinically relevant bacteria. We detected widespread CHG-tolerant bacteria (≥ 18.75 µg/mL), including opportunistic pathogens (e.g., *Pseudomonas aeruginosa*, *Stenotrophomonas maltophilia*, *Elizabethkingia miricola*), with minimum inhibitory concentrations up to 512 μg/mL. Sink drains emerged as critical hotspots, and indoor air as a potential transport mechanism. We observed indications of bacterial persistence, increased tolerance, in-situ evolution, and dissemination across MICU rooms. Molecular analyses revealed heterogeneous and largely unexplored CHG resistance mechanisms and identified resistance determinant candidates, particularly *qacEdelta1*-carrying, plasmid-borne multidrug-resistant cassettes. Our findings underscore the importance of understanding human-environment and chemical-microbe interactions to preserve chlorhexidine’s efficacy and inform infection prevention strategies. We advocate for integrated environmental management and clinical interventions under the One Health framework.

## Introduction

The emergence of resistance to antimicrobials is a critical concern worldwide, severely limiting our ability to prevent and treat infections. Chlorhexidine digluconate (CHG) has been widely used as a topical antiseptic in hospitals), both for patient skin antisepsis and for healthcare provider hand hygiene. Other clinical applications of CHG include antiseptic bathing of residents of long-term care facilities, skin preparation before invasive outpatient procedures and surgeries, and as a mouthwash for treating gingivitis. Additionally, it is extensively utilized in veterinary medicine. Chlorhexidine has broad antimicrobial activity – it is bactericidal at higher concentrations (>0.12%) and bacteriostatic at lower concentrations (0.02%-0.06%)^1^ – and it has been used for decades with little concern for resistance. While daily bathing of patients with 2% (20,000 µg/mL) CHG has served as an effective measure for prevention of healthcare-associated infections^2–4^, exposure of bacteria to sublethal concentrations of chlorhexidine can lead to reduced susceptibility to this antiseptic and other clinically relevant antibiotics (e.g., erythromycin, clindamycin, daptomycin, vancomycin, azithromycin and ciprofloxacin)^5–8^. Post-application, CHG may be shed from the skin onto indoor surfaces, resulting in sublethal concentrations of CHG in the patient’s surrounding environment. Furthermore, CHG residuals might persist in the built environment, even after undergoing degradation by heat, light, and other chemicals^9–11^.

Decreased susceptibility to CHG has been observed in various bacterial taxa (e.g., *Acinetobacter* spp*., Pseudomonas* spp*., Enterobacter* spp., and *Enterococcus* spp.), including clinically significant pathogens (e.g., *Klebsiella pneumoniae,* methicillin-resistant *Staphylococcus aureus* [MRSA]). Cross-resistance of these isolates to colistin has also been observed, raising concerns about unintended consequences of CHG application in healthcare settings^12–14^. Studies have obtained correlations between CHG susceptibility and antibiotic susceptibility in both minimum inhibitory concentrations (MICs) and minimum bactericidal concentrations (MBCs) among Gram-negative bacteria, and MBCs among Gram-positive bacteria^15^. Sporadic cases of hospital infection outbreaks caused by highly CHG-resistant *Serratia marcescens* strains have been reported^16–19^. The concern is magnified considering the inductive effect of widely used antimicrobial chemicals (e.g., triclosan, CHG) on the dissemination of antibiotic resistance genes (ARGs)^20,21^, and the possibility that mobile antibiotic resistance elements could transfer from environmental bacteria to human pathogens. In a veterinary hospital, multi-drug resistant *Serratia* isolates with similar resistance profiles were detected from animals and CHG solutions used across the hospital. Furthermore, the authors identified an IncHI2 multi-drug resistance plasmid that is shared by these *Serratia* isolates and an *Enterobacter hormaechei* strain previously recovered from the hospital environment, with the plasmid’s mobility confirmed by conjugation experiments^19^.

Generally, studies about CHG tolerance are restricted to human subjects (e.g., clinical isolates from patients, especially those on the WHO priority list of antibiotic-resistant bacteria), while little attention has been paid to the surrounding environments, which can serve as hotspots of antibiotic-resistant bacteria and hospital-acquired infections. To date, CHG susceptibility of environmental microbes has been reported only sporadically. These reports are often associated with outbreak investigations^19,22–24^, while comprehensive surveillance is lacking. Comprehensive CHG surveillance is partly impeded by the lack of standardized terms, including resistance, tolerance, and reduced susceptibility. In this study, we defined chlorhexidine resistance using epidemiological cut-off values proposed previously (Table S1)^12,25,26^, due to the lack of standardized clinical breakpoints for chlorhexidine^27^. The values were defined as the upper limit of the normal MIC distribution for a given antimicrobial and wild-type species. Clinical implications of these values are unclear, but they are nevertheless helpful in resistance monitoring. In cases where epidemiological cut-offs are also unknown, we interchangeably used the terms tolerance and reduced susceptibility as general descriptors. In this study, chlorhexidine digluconate was used for MIC measurements, and chlorhexidine MICs were used as a general term for MICs reported by previous studies where the chemical was not explicitly stated. In cases where the taxonomy or the epidemiological cut-off values were unknown, for uniformity reasons, we described MICs ≥ 128 µg/mL as high CHG MICs, MICs of 512 µg/mL as the highest CHG MICs, and bacteria that grew at 18.75 µg/mL CHG as CHG tolerant^3^.

In this study, we conducted the most comprehensive exploration of CHG tolerance in MICU environments to date, examining chemical, microbial, and molecular aspects. Through controlled microcosm experiments, we gained insights into the fate of CHG on surfaces and the factors influencing its persistence. In subsequent field surveys, we identified environmental hotspots harboring CHG-tolerant, opportunistic, persistent, and disseminating bacteria. Additionally, we identified candidate and novel ARGs and mobile genetic cassettes potentially associated with CHG tolerance.

## Results

### Residual chlorhexidine persists on surfaces under disinfection and cleaning in microcosm experiments

CHG in the environment can degrade naturally due to environmental factors such as heat and light. Additionally, physical removal through cleaning or reactions with cleaning products may decrease residual surface concentrations. We investigated CHG persistence on three common hospital surface types - laminate wood, plastic (high-density polyethylene; HDPE), and metal (stainless steel) - under six cleaning practices. These practices included no cleaning, cleaning with water, and disinfecting with ethanol, bleach, peracetic acid, or benzalkonium chloride (Fig. S1).

While the concentration of CHG decreased over time, it persisted on indoor surfaces over 24 h (Fig. 1a, Table S2). The persistence patterns of CHG residual were impacted by surface material and cleaning practice. We first investigated the immediate impact of wiping a surface on CHG persistence by comparing the concentrations at time 0 (i.e., the amount applied) and 10 min (i.e., 0.17 h). Among the cleaning practices, wiping with peracetic acid had the largest immediate impact, showing significant differences on plastic and laminate wood surfaces (*p* < 0.02) and approaching significance on metal surfaces (*p* = 0.056). This is supported by previous evidence that CHG can be inactivated at a low pH and in the presence of anionic molecules^28–30^. In contrast, none of the other disinfectants tested are known to interact with CHG. Benzalkonium chloride and ethanol have been shown to have synergistic or additive antimicrobial properties when applied with CHG, indicating that their co-occurrence will not cause chlorhexidine degradation^31–35^. This is concordant with our observation that CHG concentration is not significantly affected by benzalkonium chloride or ethanol application.

**Fig. 1.**
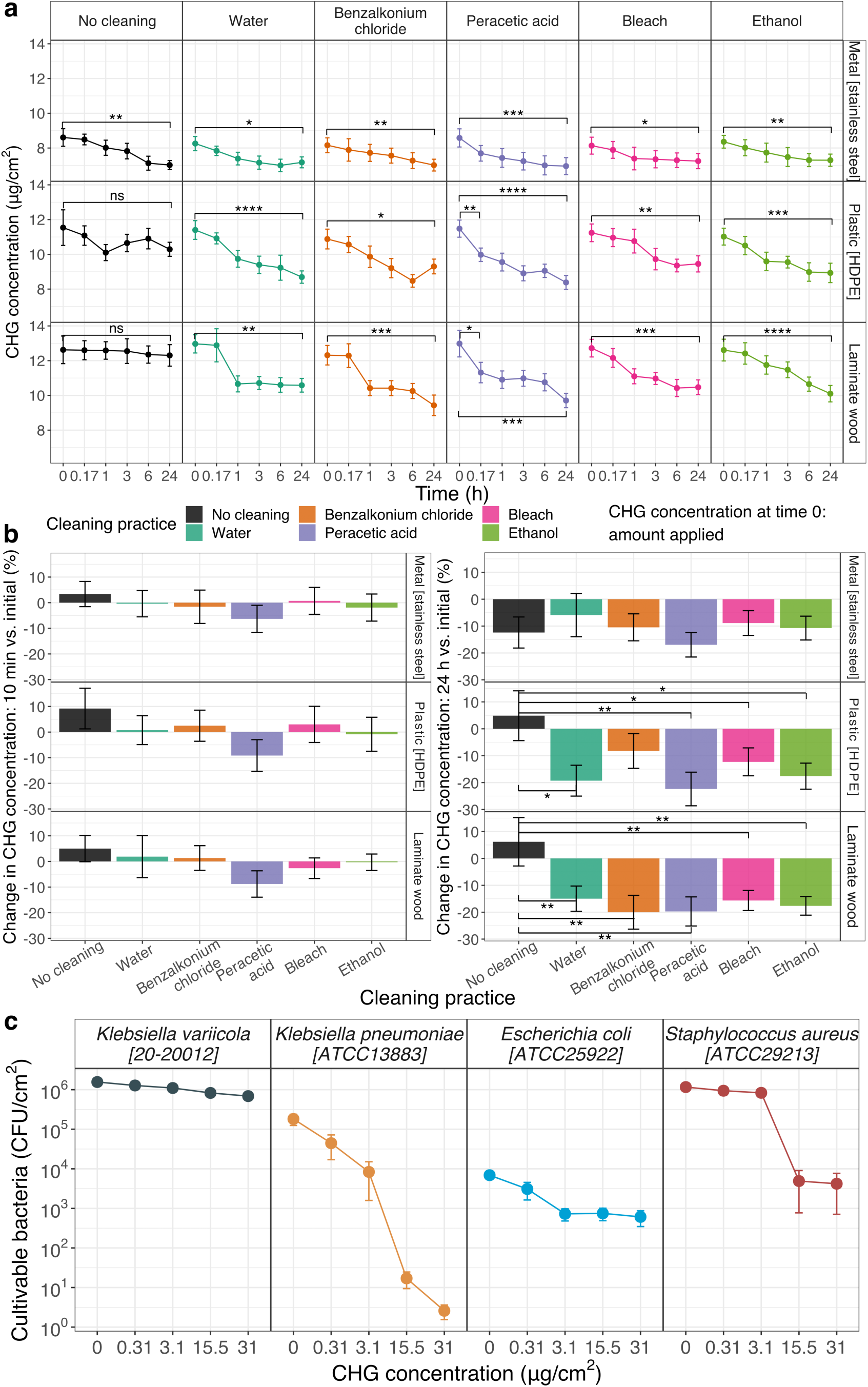
Residual CHG persists on surfaces despite cleaning and disinfection, with concentrations decreasing to sublethal levels for clinically relevant bacteria. **a)** The change patterns of CHG concentrations on surfaces over time in response to cleaning and disinfection. Significance was determined by paired *t*-tests with the alternative hypothesis that the CHG concentration at 0 h was greater than that at 0.17 h (or 24 h). For the immediate impact (0 h vs 0.17 h), only groups with significant differences were labeled for visual clarity. Recovery rates in sampling and measurement were accounted for in the calculations. **b)** Immediate and long-term reduction effects of cleaning practices across surface types. Significance was determined by pairwise *t*-tests relative to the “no cleaning” group or the “cleaning with water” group, with the alternative hypothesis that the tested groups had a greater reduction effect than the reference group. Multi-test *p*-values were adjusted using the Benjamini-Hochberg method. Only significant associations were labeled; thus, only cleaning plastic and laminate wood showed a significantly greater reduction effect than no cleaning. **c)** Clinically relevant bacteria can survive 24-hour exposure to CHG residual concentrations on plastic surfaces.

Although CHG concentration significantly decreased after 24 h on all cleaned surfaces, it still persisted on indoor surfaces with residual concentrations ranging from 7.0 to 12.3 μg/cm^2^ (Fig. 1a). When cleaning was not conducted, CHG concentrations remained stable on laminate wood and plastic surfaces, whereas a significant decrease was observed on metal surfaces (*p* = 0.005). While all cleaning methods induced significant concentration reductions after 24 h (Fig. 1a), only cleaning plastic and laminate wood showed a significantly greater reduction effect than no cleaning (*p* < 0.05), whereas disinfecting did not statistically differ from cleaning with water (Fig. 1b). This suggests that, for all tested surface types, potential interactions between disinfecting chemicals and CHG do not exhibit a notably greater reduction effect than the physical removal of CHG during cleaning. Our findings align with previous indications that chlorhexidine degradation might be catalyzed by metals and that chlorhexidine is unstable under acidic and alkaline conditions (pH 3.5-6.5 and pH > 8.5)^36^.

### Clinically relevant bacteria can survive 24 h low-dose chlorhexidine exposure on surfaces

We examined the survival of four clinically relevant bacterial isolates (*Escherichia coli* [ATCC 25922], *Klebsiella pneumoniae* [ATCC 13883], *Klebsiella variicola* [20-20012]^37^, *Staphylococcus aureus* [ATCC 29213]) on plastic surfaces after exposure to CHG for 24 h (Fig. 1c). Plastic was selected as a representative surface because it is a common material of equipment in the immediate vicinity of patients and healthcare workers (e.g., bedrail, nurse call button, keyboard, mouse). Five concentrations of CHG (31.00, 15.50, 3.10, 0.31, and 0 μg/cm^2^) were tested to encompass the range persisting on surfaces after cleaning or disinfection.

After 24 h, all species of bacteria survived on CHG-treated surfaces across all tested microbial densities and CHG concentrations (2.58 – 1.28×10^6^ CFU/cm^2^, log_10_ reduction ranging from 0.08 to 4.87). Although fewer *Klebsiella pneumoniae* CFUs survived at 15.5 and 31 μg/cm^2^ CHG, an estimation from the data trend suggests that >17 CFU/cm^2^ could survive at CHG concentrations of 8.4-11.5 μg/cm^2^ (range of CHG persisting on plastic surfaces). As previous studies have linked exposure to sublethal concentrations of chlorhexidine with reduced susceptibility to chlorhexidine and clinical antibiotics such as vancomycin and daptomycin^6–8,38^, these results raise concerns about potential development of antimicrobial resistance in bacteria on surfaces that possibly harbor CHG residues (e.g., hospital environments where patients are bathed with CHG).

### Chlorhexidine-tolerant bacteria are widespread in MICU environments with sink drains being critical hotspots

Our microcosm experiments indicated that the environment may serve as a hotspot for CHG-tolerant bacteria. To determine how widespread CHG tolerance is *in situ*, we further conducted a field survey of environmental locations in a MICU (Methods, Fig. S2). Cultivation yielded a total of 1415 isolates of 345 different phenotypic morphologies, of which 306 isolates (21.6%) were confirmed to grow at 18.75 µg/mL CHG at 25 °C (Fig. S3). In this study, 18.75 µg/mL was used as the threshold to describe CHG tolerance of bacteria^3^. Additionally, 203 isolates (14.3%) were suspected to grow; due to colony proximity on the agar, it was difficult to determine whether they grew independently or were protected by neighboring colonies. All 509 isolates (36.0%) underwent CHG MIC testing.

We found that unit area surface bioburden in the MICU was significantly associated with touch and cleaning frequencies (see Methods for definitions of touch frequencies), and the overall trend was consistent with a previous study^39^. The rarely cleaned doorsills had significantly higher bioburden than high-touch bedrails and nurse call buttons, followed by medium-touch keyboards (unpaired *t*-tests, log_10_ scale) (Fig. 2a). This trend was also consistent for the absolute bioburden in the entirety of the surfaces (Fig. S4). The absolute bioburden on low-touch light switches was the lowest. However, they exhibited the highest unit area bioburden among dry surfaces. It is worth noting that the switches had the smallest surface area (Table S3), which may have contributed to greater uncertainty in calculating the unit area bioburden. No effect of sampling seasons and patient room isolation status (contact isolation or not) was observed (Fig. S5).

**Fig. 2.**
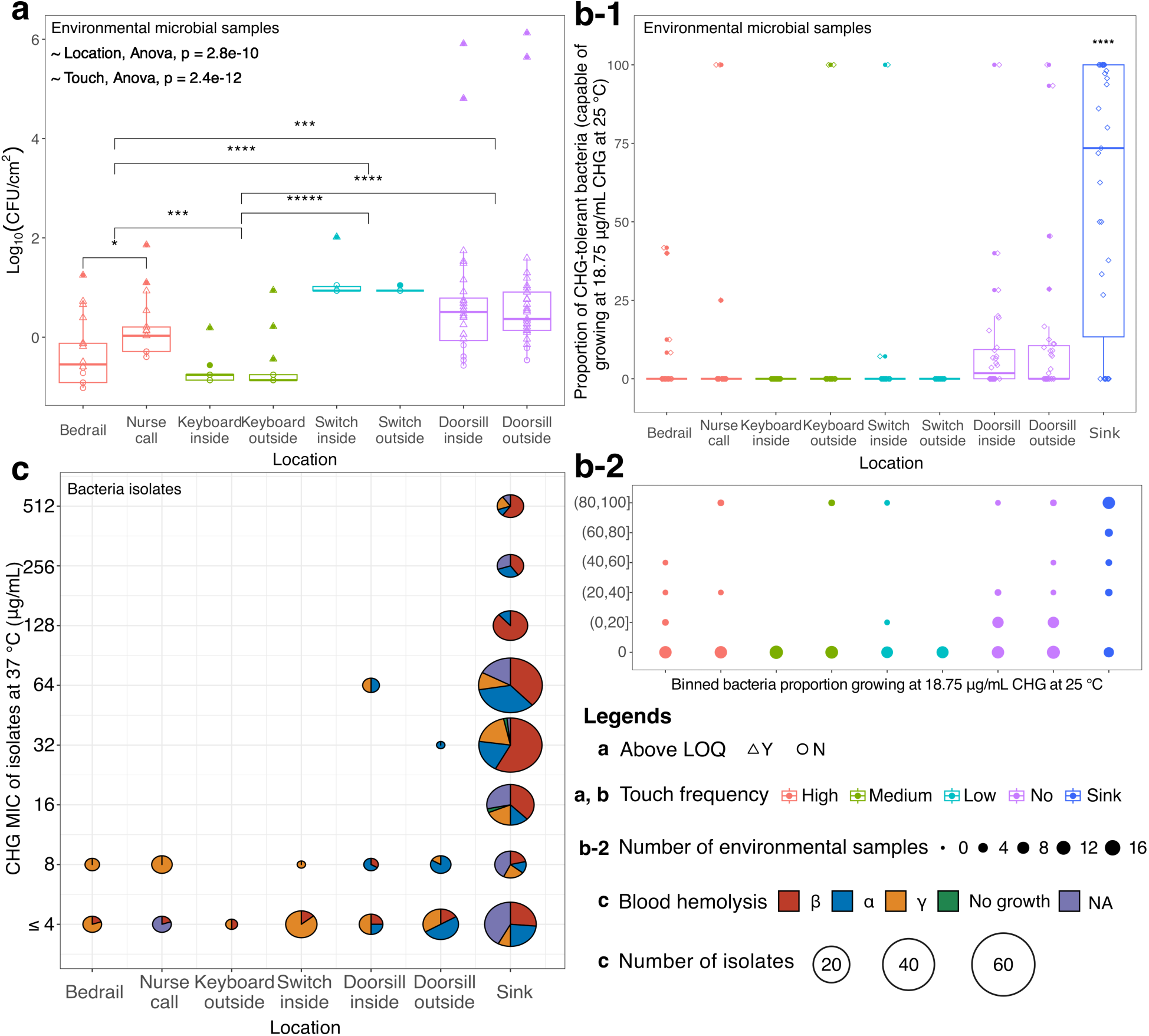
Chlorhexidine-tolerant bacteria are widespread in MICU environments with sink drains being critical hotspots. **a)** Unit area bioburden of bacteria across different touch frequencies on dry surfaces (cultivation temperature: 25 °C). CFU: colony forming unit. To avoid mis-interpreting data with small colony counts, we set the limit of quantification (LOQ) of our culture efforts as colony counts = 3. Values below this threshold were recorded as half of LOQ (i.e., 1.5). **b)** Proportion of bacteria tolerant to 18.75 µg/mL CHG (described as CHG tolerant in this study) at 25 °C across different touch frequencies on dry surfaces and sink. The proportion of sink samples was significantly higher than that of all the other touch groups according to unpaired *t*-tests (p ≤ 0.0001 for all comparisons). In both a) and b), only significant comparisons were labeled for visual clarity. **c)** Of the bacterial isolates growing at 18.75 µg/mL CHG at 25 °C, distribution of their CHG MICs at 37 °C and the corresponding blood hemolysis status. Nurse call button is abbreviated as nurse call in the figure.

Despite CHG primarily being applied to patient skin, CHG tolerance was identified in multiple spatially distinct locations within the MICU. Most bacteria on dry surfaces were not CHG tolerant at 25°C (Fig. 2b), defined as the ability to grow at 18.75 µg/mL CHG. The percentages of bacteria exhibiting tolerance did not significantly differ among dry surfaces, with mean percentages ranging from 0% (keyboards in patient rooms, light switches in the hallway) to 13.2% (nurse call buttons). Potential pathogens (as indicated by β hemolysis on blood agar at 37 °C) were detected across all environmental locations^40^. However, doorsills were the only dry surfaces where CHG tolerance at 37 °C was detected (Fig. 2c, 32 µg/mL - one isolate, 64 µg/mL - four isolates). There was also a slight trend of a subpopulation of doorsill samples harboring high proportions of CHG tolerant bacteria (3 out of 59 samples had percentages >90%).

Sink drains were critical hotspots for absolute bioburden (Fig. S4), CHG tolerance (Fig. 2b-c), and potential pathogens (Fig. 2c). The absolute bioburden and proportion of CHG-tolerant bacteria in sink drains were significantly higher than on all dry surfaces (*p* ≤ 0.0001). Moreover, 37 bacterial isolates with high CHG MICs (≥ 128 µg/mL) were detected from sink drains in patient rooms, medication rooms, and MICU hallways (128 µg/mL - 17 isolates, 256 µg/mL - 10 isolates, 512 µg/mL - 10 isolates) (Fig. 2c). Of the CHG-tolerant bacteria at 25 °C from the sink drains, 70.6% (127 out of 180 isolates) were β-hemolytic (a highly predictive indication of pathogenicity^40^), including these with high MICs (24 out of 29 isolates, Fig. 2c).

### Opportunistic pathogens and high CHG MIC bacteria were detected in MICU environments with increased tolerance and potential for persistence

To further investigate their potential clinical importance and genomic resistance mechanisms, we performed whole-genome sequencing on 63 isolates (Fig. S6, Table S4). The isolates were randomly selected, with efforts to ensure coverage of various colony morphologies and MICs.

Additionally, comparisons were prioritized by selecting isolates with different MICs but the same colony morphology. Among these, the three most represented species are opportunistic pathogens: *Stenotrophomonas maltophilia* (n = 19), *Elizabethkingia miricola* (n = 7), and *Pseudomonas aeruginosa* (n = 6). Isolates with high CHG MICs were detected among these species, with the highest values being 128, 512, and 128 µg/mL, respectively. Notably, all these isolates were recovered from sink drains located in diverse spaces across patient rooms, medication rooms, and hallways, except for one *S. maltophilia* isolate from a doorsill in the hallway. Additionally, we detected two *Acinetobacter radioresistens* isolates from the doorsill of a patient room, both carrying the carbapenemase gene *bla*_OXA-23_. While there were no patient infections or colonization by *S. maltophilia* or *E. miricola* at the sampled MICU during the sampling sessions, these species have previously been reported to develop multi-drug resistance and be associated with nosocomial and community-acquired infections. Specifically, *S. maltophilia* is associated with respiratory infections^41^. *E. miricola* has been reported to associate with a variety of infections including bacteremia, pulmonary abscess, urinary tract infection, catheter-associated bacteremia in a hemodialysis patient, and a fatal intracranial infection^42–49^.

*Elizabethkingia* has also been implicated in healthcare-associated outbreaks of the built healthcare environment^50^. *P. aeruginosa* can cause pneumonia and bacteremia, along with urinary tract and wound infections^51^. It is one of the three species ranked first priority on the World Health Organization (WHO) list of antibiotic-resistant bacteria^52^ and a pathogen of major concern for nosocomial infections^53^. Although rare, *A. radioresistens* can cause infections in the lungs, blood, wounds, or urinary tracts, particularly in hospital settings^54–57^. More importantly, it has been identified as a potential disseminator of ARGs, transferring the *bla*_OXA-23_ gene to *Acinetobacter baumannii*^58^, another first-priority pathogen on the WHO list.

Moreover, we identified five genomically different isolates belonging to novel species according to multiple identification algorithms (KmerFinder, PhyloPhlAn, autoMLST, and MiGA), all of which were recovered from sink drains and three of which are potential pathogens (β-hemolysis on blood agar^40^) harboring multiple copies of multidrug-resistant ARGs. Three can be identified to genus level (*g_Stenotrophomonas*, *g_Achromobacter*, *g_Cupriavidus*), while the other two can only be identified to family level (*f_Rhizobiaceae*, *f_Xanthomonadaceae*).

Interestingly, in this subgroup (63 out of 1415 isolates), we observed potential evidence of bacterial persistence and in situ evolution of opportunistic pathogens in sink drains, as well as their potential dissemination across multiple MICU rooms. In both sampling events (February and July), we recovered the same strains (ANI ≥ 99.99%^59,60^) of *P. aeruginosa* and *S. maltophilia* from the same sink drains across three patient rooms (PT_3, PT_4, PT_8, Fig. 3a). Observation of the same strain in the same location at two time points suggests that it persisted in sink drains over the course of half a year. Notably, the original *P. aeruginosa* isolate and one of the subsequent isolates carried type-1 *qacEdelta1*-carrying plasmids and exhibited high CHG MICs (128 µg/mL, above *P. aeruginosa*’s epidemiological breakpoint 50 µg/mL, Fig. 6, Table S1). However, a second subsequent isolate, while still meeting the identity threshold for belonging to the same strain, did not contain the *qacEdelta1* gene and had an MIC of 32 µg/mL. This observation suggests that the ARG-carrying plasmid can be lost, potentially under reduced selective pressure, and that carrying *qacEdelta1* may associate with CHG resistance in *P. aeruginosa* (Table S1). For *S. maltophilia*, we observed signs of increased CHG tolerance and intra-species/inter-strain competition during the persistence. In the PT_3 sink drain, the MIC of two intra-strain isolates increased from 16 to 64 µg/mL between the first and second sampling events. In the PT_8 sink drain, we identified two inter-strain isolates with MICs of 4 µg/mL (S60) and 32 µg/mL (S10) during the first sampling event, and one isolate with an MIC of 64 µg/mL, belonging to the same strain as S10, during the second event. Furthermore, in sink drains of multiple rooms, we detected the same strains of *E. miricola* (MD_3 and PT_10, PT_2 and PT_7) and *S. maltophilia* (PT_7 and PT_8) as well as the same species of *P. aeruginosa* and *Delftia tsuruhatensis* (Fig. 3b, Fig. S7). Additionally, we found that *S. maltophilia* and *P. aeruginosa* cohabitated the same sink drain in two patient rooms (PT_5, PT_7). These organisms can be co-colonizers in the respiratory tract of cystic fibrosis patients^41^, indicating the possibility of contracting both pathogens from the same environmental location^61^. It should be noted that these findings are based on a limited number of isolates due to a tradeoff with taxa breadth. Nevertheless, they offer guidance for future studies with larger sample sizes, potentially focusing on specific species.

**Fig. 3.**
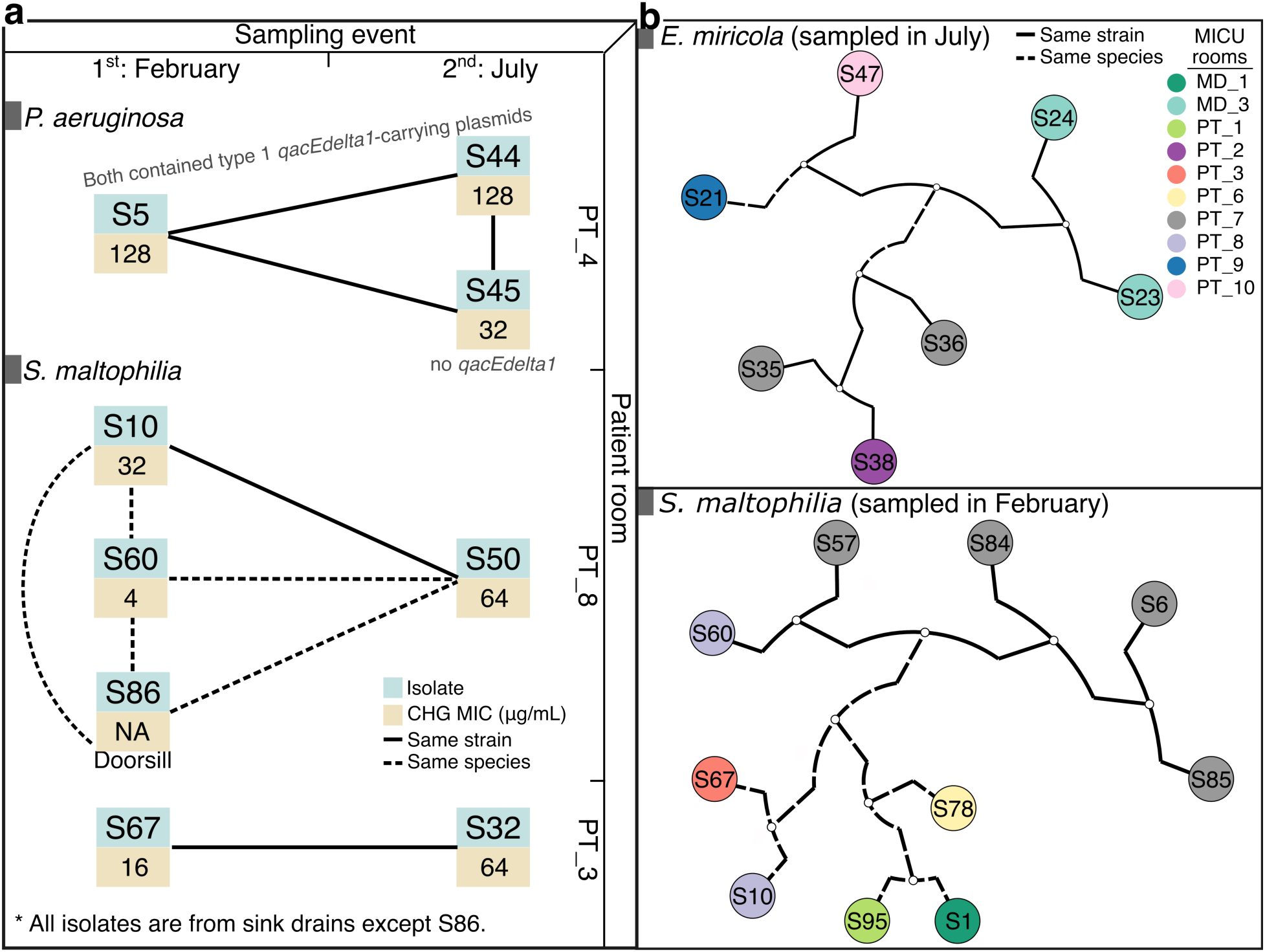
Indications of a) bacterial persistence and in situ evolution of opportunistic pathogens in sink drains, as well as b) their potential dissemination across multiple MICU rooms. Patient rooms were deidentified. See Fig. S7 for additional species with multiple isolates detected in several MICU rooms during the same sampling event.

Of the 37 high-CHG MIC isolates in the cultivation dataset, 19 were sequenced. Among these, we found three *S. maltophilia*, two *P. aeruginosa*, seven *E. miricola*, two *Cupriavidus metallidurans*, and two *D. tsuruhatensis* isolates. The remaining three isolates had ambiguous species assignments possibly belonging to *D. tsuruhatensis*, *Delftia acidovorans*, or *S. maltophilia*. While *S. maltophilia*, *P. aeruginosa*, and *E. miricola* are more often recognized in clinical cultures, *C. metallidurans*, *D. tsuruhatensis*, and *D. acidovorans* are gaining attention as emerging opportunistic pathogens^62–67^.

### Molecular mechanisms conferring chlorhexidine resistance are heterogeneous and remain largely undiscovered

Across high-CHG-MIC species, antibiotic efflux was the most prevalent and abundant identified resistance mechanism (Fig. 4a). *P. aeruginosa* harbored a significantly greater quantity and variety of ARGs than other species. However, while *C. metallidurans* harbored fewer ARGs, most of its ARGs were mobile (6 out of 8 on average). ARG profiles of high-CHG-MIC species demonstrated heterogeneity both within and across species (Fig. 4b-c). We detected several ARGs falling into Q1 of the health risk index ranking^68^ in ≥ 2 species (out of 7), including *sme* family, *sul1*, and *adeF*. Particularly, *adeF* was present in all high-CHG-MIC isolates except *P. aeruginosa*.

**Fig. 4.**
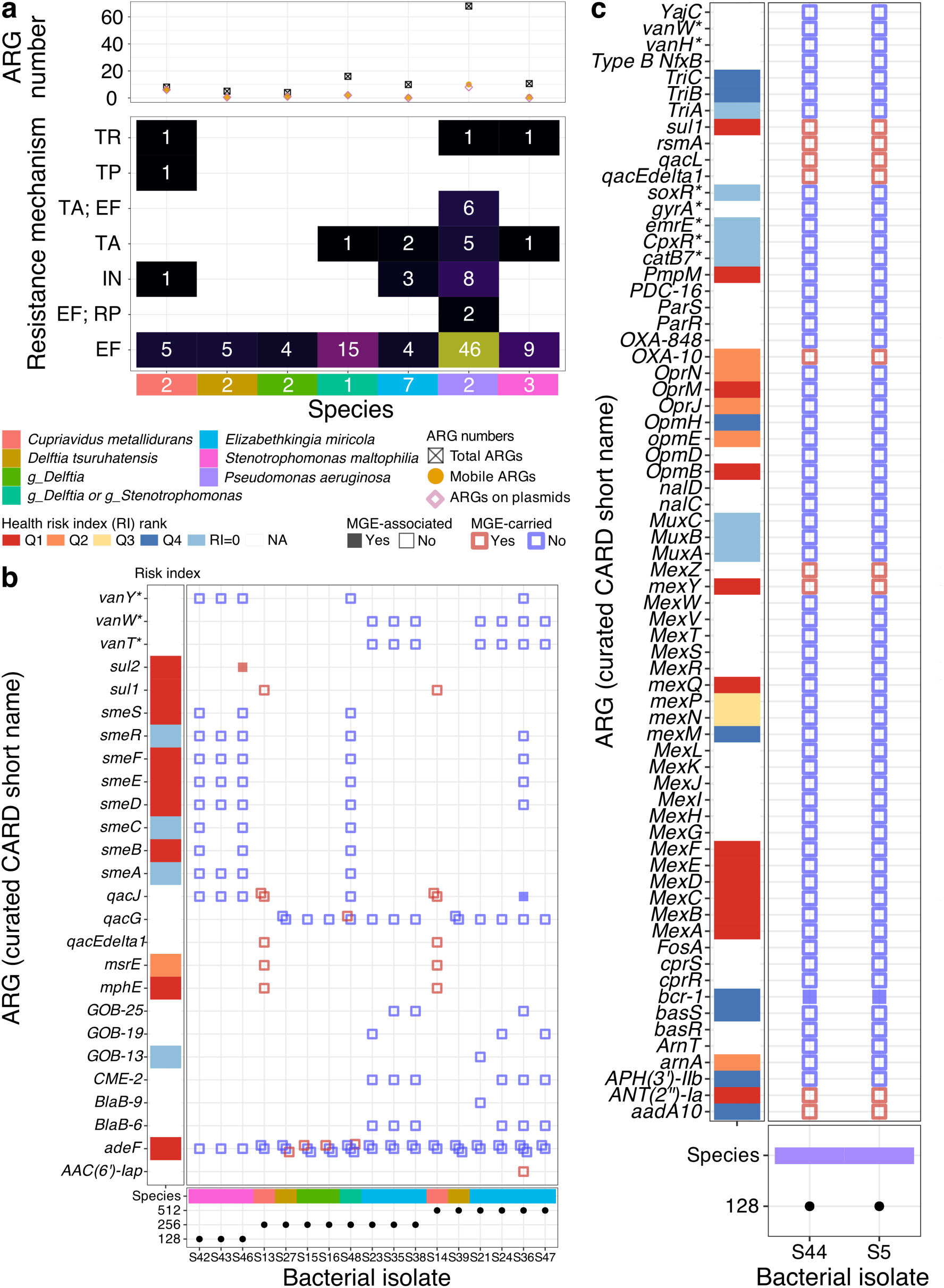
Antibiotic resistance mechanism and gene profiles of the sequenced high-CHG-MIC isolates (MICs ≥ 128 µg/mL). **a)** Distribution of antibiotic resistance mechanisms and genes across different species. ARG numbers: Error bars represent the mean standard error. Resistance mechanisms: Numbers in the main heatmap represent the average ARG counts for each mechanism; those in the marginal heat blocks indicate the number of high-CHG-MIC isolates per species. Antibiotic resistance gene profiles of high-CHG-MIC isolates: **b)** all species except *Pseudomonas aeruginosa*, **c** *Pseudomonas aeruginosa*. Each square in the dot plots represents one ARG copy. ARGs were named according to the curated CARD short names (output of Resistance Gene Identifier, Methods)^76^. ARGs located within MGEs were classified as MGE-carried; those situated within a 5 kb proximity to an MGE were considered MGE-associated; both were deemed mobile. Health risk index ranks from Zhang et al. were used to prioritize ARGs^68^. Abbreviations: **a)** EF: efflux, IN: inactivation, RP: reduced permeability, TR: target replacement, TP: target protection, TA: target alteration; **b)** *vanY\**: *vanY* gene in *vanM* cluster, *vanW\**: *vanW* gene in *vanG* cluster, *vanT\**: *vanT* gene in *vanG* cluster; **c** *vanH\**: *vanH* gene in *vanB* cluster, *gyrA\**: *Pseudomonas aeruginosa gyrA* conferring resistance to fluoroquinolones, *emrE\**: *Pseudomonas aeruginosa emrE*, *CpxR\**: *Pseudomonas aeruginosa CpxR*, *catB7\**: *Pseudomonas aeruginosa catB7*, *soxR\**: *Pseudomonas aeruginosa soxR*; **legend)** *g_ Delftia*: ambiguous species *Delftia tsuruhatensis* or *Delftia acidovorans*, *g_Delftia* or *g_Stenotrophomonas*: ambiguous species *Delftia tsuruhatensis*, *Delftia acidovorans* or *Stenotrophomonas maltophilia*.

Although many ARGs were identified, only two known to reduce susceptibility to CHG, *qacEdelta1*^69–71^ and *norA*^72–75^, were observed in 63 sequenced isolates (Table S4). Among the high-CHG-MIC isolates, only *qacEdelta1* was detected, and this was limited to four out of 19 such isolates belonging to two species (*C*. *metallidurans* and *P*. *aeruginosa*, Fig. 4b-c). In addition, two *Acinetobacter radioresistens* isolates sharing the same ARG profile exhibited ≥ 16-fold CHG MICs difference (Fig. 5c). These, in combination, indicate that novel ARGs or pathways conferring CHG resistance remain to be discovered.

**Fig. 5.**
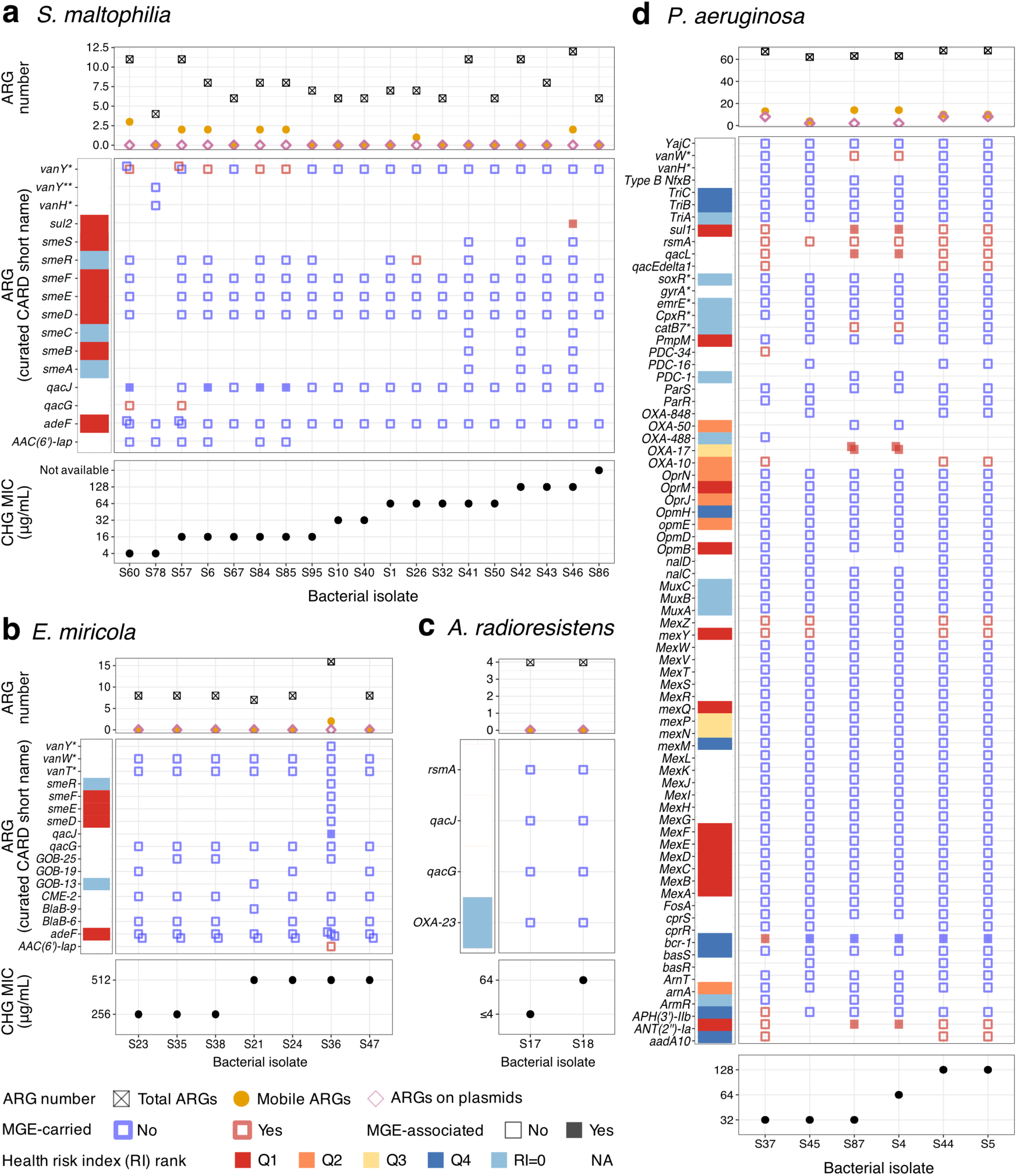
ARG profiles of isolates with varying CHG MICs, including ARG mobility and health risk index for: a) *Stenotrophomonas maltophilia*, b) *Elizabethkingia miricola*, c) *Acinetobacter radioresistens*, and d) *Pseudomonas aeruginosa*. Each square in the dot plots represents one ARG copy. ARGs were named according to the curated CARD short names (output of Resistance Gene Identifier, Methods)^76^. ARGs located within MGEs were classified as MGE-carried; those situated within a 5 kb proximity to an MGE were considered MGE-associated; both were deemed mobile. Health risk index ranks from Zhang et al. were used to prioritize ARGs^68^. Abbreviations: a) *vanY*\*: *vanY* in *vanM* cluster, *vanY*\*\*: *vanY* in *vanA* cluster, *vanH*\*: *vanH* in *vanO* cluster; b) *vanW*\*: *vanW* gene in *vanG* cluster, *vanT*\*: *vanT* gene in *vanG* cluster.

To further investigate the genetic characteristics of clinically important bacteria regarding CHG tolerance, we examined the presence, types, and differential abundances of ARGs and mobile ARGs with varied CHG MICs in *S. maltophilia*, *E. miricola*, *A. radioresistens*, and *P. aeruginosa* (Fig. 5). Among these species, *P. aeruginosa* was the only one found to carry a known CHG-ARG, *qacEdelta1*, which was present in three out of the six isolates. Specifically, two isolates had MICs of 128 µg/mL and one had an MIC of 32 µg/mL. In all cases, the gene was located on a plasmid, indicating potential for horizontal gene transfer. Although no known CHG ARG was predicted in the other two species, upon qualitative inspection, we identified ARGs potentially associated with CHG tolerance. These included *sul2*, *smeA/B/C/S* for *S. maltophilia*, and *smeD/E/F/R*, *qacJ*, *AAC(6’)-Iap* for *E. miricola*. Future research could interrogate these gene candidates, especially the Q1 health risk ARGs^68^ *sul2* and *smeB/D/E/F/S*, by employing more quantitative and systematic surveys along with experimental validations. The taxon *E. miricola* may be associated with high CHG MICs, as all isolates exhibited MICs of 256 µg/mL or higher. Particularly, isolate S36 had an MIC of 512 µg/mL. Compared to other *E. miricola* isolates, it contained more types and quantities of ARGs, and more critically two unique mobile ARGs (*qacJ* and *AAC(6’)-Iap*). *P. aeruginosa* exhibited significantly higher quantities of ARGs and mobile ARGs compared to most species in the dataset (Table S4) and consistently ranked in the top three for all three indices. Notably, several isolates warrant further attention: isolate S30, with an ambiguous species assignment between *Citrobacter amalonaticus* and *Citrobacter freundii*; S25, belonging to a novel species of genus *Cupriavidus*; and S13 and S14, identified as *<u>Cupriavidus metallidurans</u>*, all contained a high number of ARGs or mobile ARGs. However, we could not establish statistical correlations between CHG MICs and the quantities of ARGs, mobile ARGs, or plasmid-borne ARGs due to the broad scope of the study and the limited number of sequenced isolates.

### *qacEdelta1*-carrying multidrug-resistant plasmids are a potential concern

*qacEdelta1* was detected exclusively on plasmids in six isolates (Fig. 6, Table S5). Each isolate harbored one *qacEdelta1*-carrying plasmid. All isolates were recovered from sink drains, with S30 originating from a staff-only restroom and the remaining coming from patient rooms. Notably, all six *qacEdelta1*-carrying plasmid scaffolds had ≥ 3 ARGs regardless of species, five of which carried the highest number of ARGs among all plasmid scaffolds in the sequenced isolates (six ARGs in one plasmid scaffold). Another notable feature shared by these plasmids was the colocalization of *qacEdelta1* and *sul1*, arrayed next to each other as a cassette. Importantly, *sul1*, a Q1 health risk ARG, confers resistance to sulfonamide antibiotics and its proximity to *qacEdelta1* merits attention.

**Fig. 6.**
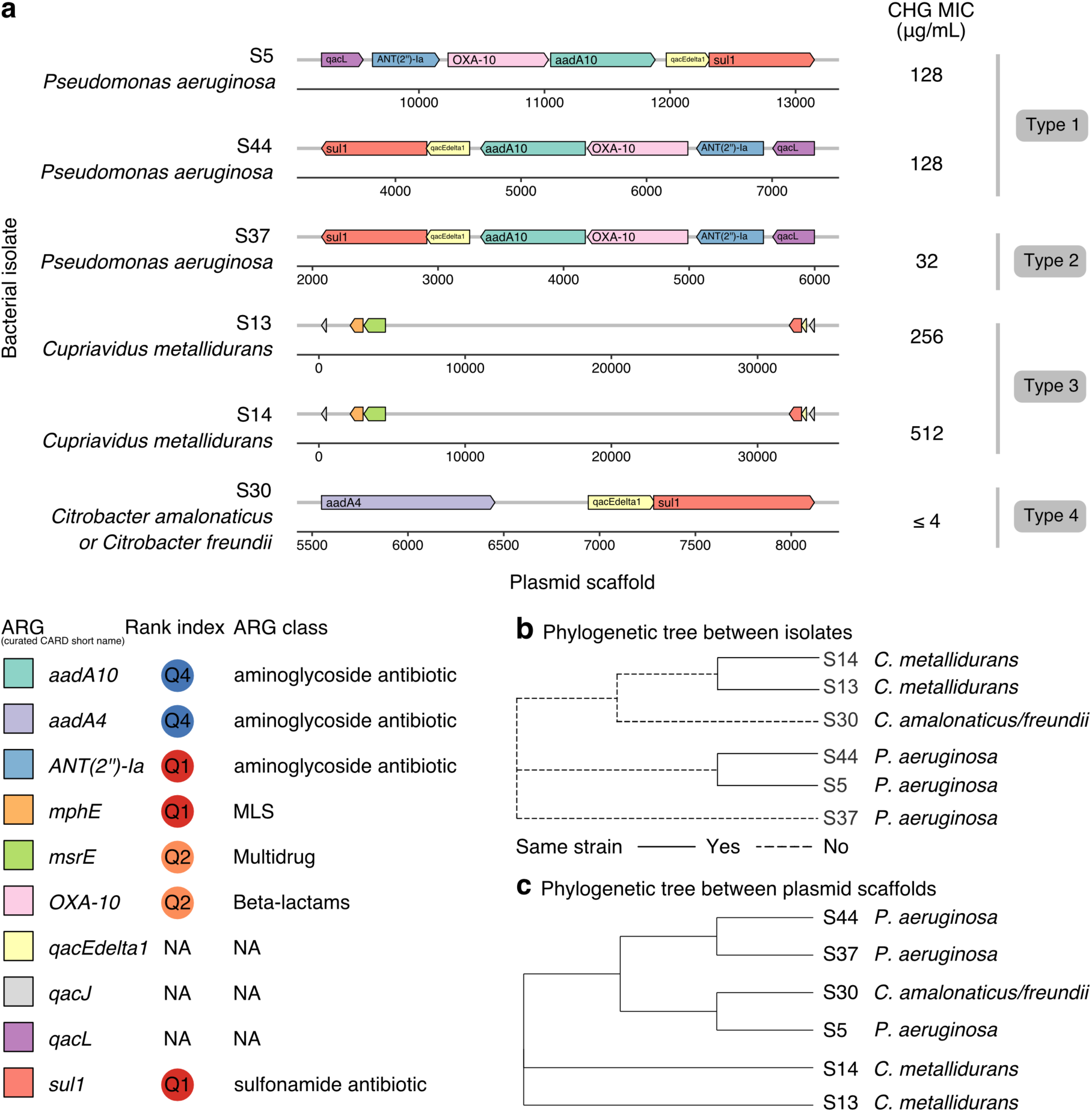
*qacEdelta1*-carrying multidrug-resistant plasmids are of potential concern, particularly type-1 and type-3 plasmid scaffolds, which are possibly associated with high CHG tolerance. **a)** Colocalization of ARGs on the plasmid scaffolds. **b)** Phylogenetic relationship between the isolates. **c)** Phylogenetic relationship between the plasmid scaffolds. All isolates were recovered from sink drains. S30 was from a staff-only restroom, and the other isolates were from patient rooms. See Table S5 for more information. ARGs were named according to the curated CARD short names (output of Resistance Gene Identifier, Methods)^76^. Health risk index ranks from Zhang et al. were used to prioritize ARGs^68^. MLS: macrolides, lincosamides and streptogramins.

The *qacEdelta1*-carrying plasmid scaffolds could be classified into four types according to blastn alignments^77^, and type-1 and type-3 plasmid scaffolds were possibly associated with high CHG tolerance. Type 1 contained two identical plasmid scaffolds in two isolates of the same *P. aeruginosa* strain (S5, S44). Type 2 comprised a plasmid scaffold from another *P. aeruginosa* strain (S37) that was closely related to type 1 with 8 bases unaligned. Type 3 was composed of the other two identical plasmid scaffolds in two isolates of the same *C. metallidurans* strain (S13, S14). Type 4 held a different plasmid scaffold in an isolate with an ambiguous species assignment between *Citrobacter amalonaticus* and *Citrobacter freundii* (S30) which shared common regions with the other plasmids. It is worth highlighting that the top BLAST hits for type-1 plasmid scaffolds (query coverage: 100%, identity ≥ 99.99%, database: nr/nt) matched two plasmids from clinical *P. aeruginosa* strains - one from a bloodstream infection and the other from sputum - both collected in Chicago (Table S5). While the two type-1 plasmid scaffolds shared an identical sequence alignment in the two *P. aeruginosa* strain-matched isolates, S5 and S44, they were positioned in opposite directions and were not the most closely related phylogenetically (Fig. 6). This suggests that carriage of this ARG cassette is dynamic and large-scale genomic changes like inversions might occur. Nevertheless, they still manifest the same MIC (128 µg/mL), indicating that the inversion did not disrupt the function of CHG tolerance. Notably, the strain of S5 and S44 probably persisted in sink drains over half a year (Fig. 3a). In comparison, plasmid scaffolds of S5 - S30 and S44 - S37 were more closely related, respectively. S44 and S37 were two different strains of *P. aeruginosa*, while S5 and S30 belonged to different orders. This suggests that the *qacEdelta1*-carrying plasmid could be transferred in situ not only between environmental bacteria of the same species but also between those that are more evolutionarily distant.

To the best of our knowledge, this study is the first to report the presence of the *qacEdelta1* gene on a plasmid in *C. metallidurans*. *C. metallidurans* was traditionally considered non-pathogenic until recent case reports suggested otherwise. In 2005, this bacterium was identified in the respiratory secretions of cystic fibrosis patients, though its pathogenic effects were not understood at the time^78^. In 2011, a case of invasive nosocomial septicemia caused by *C. metallidurans* was reported following surgery^62^. Furthermore, in 2015, four cases of catheter-related bacteremia due to this bacterium were documented^79^. The growing clinical importance, combined with its detection in MICU patient room environments, high CHG tolerance, and carriage of a concerning plasmid, implicate the need for further investigations.

## Discussion

Although CHG is used primarily on patient skin, it could be transferred to hospital environments. Once there, CHG likely persists on indoor surfaces despite surface cleaning and disinfection, with its concentration decreasing to sublethal levels for some clinically relevant bacteria, as demonstrated by our microcosm experiments. This finding was further corroborated by an in-situ field survey, which revealed widespread CHG tolerance in MICU environments. Notably, we emphasized sink drains as critical hotspots, corroborating previous findings. Further, based on the isolation of CHG-tolerant bacteria from doorsills, we urge more attention to indoor air as transport mechanisms for resistant organisms. We observed potential evidence of bacterial persistence, increased tolerance, and in-situ evolution of opportunistic pathogens in sink drains, as well as dissemination across multiple MICU rooms. *S. maltophilia*, *E. miricola*, and *P. aeruginosa* were highly represented species in the sequenced subset. Through our broad-scope study, we found that the molecular mechanisms conferring chlorhexidine resistance were heterogeneous and largely undiscovered. Nevertheless, we identified ARG candidates that warrant future investigation, particularly two types of *qacEdelta1*-carrying, plasmid-borne multidrug-resistant cassettes. These cassettes are potentially concerning for their role in pathogen persistence, the development and dissemination of CHG resistance, and broader antibiotic resistance.

The ways in which humans interact with their environment, including touch and cleaning, along with characteristics like surface material, jointly impact the fate of chemicals applied to humans. Subsequently, these factors influence the trajectory of microbial ecology and evolution and in return affect our exposome^80^. We identified sink drains and air as environmental hotspots for CHG tolerance and opportunistic pathogens. Consistent with previous observations^81–83^, sink drains were confirmed as critical reservoirs. CHG-tolerant and/or opportunistic bacteria hosted in sink drains, likely within biofilms, could persist for at least half a year, adaptively evolving and increasing their CHG tolerance, and potentially disseminating^84^. We hypothesize that water availability plays an important role, driving the in-situ microbial survival and persistence as well as antimicrobial adaptation and evolution in the built environment (e.g., sink drains). It may exert a stronger impact than other factors such as touch frequency and surface material^85,86^. Building on previous evidence that microbes can be transmitted from sinks to patients^61^, our findings further emphasize the importance of sinks as possible sources of healthcare-associated infections and antimicrobial resistance, supporting the waterless ICU initiative^87^.

Since biomass on doorsills reflects what is circulating in the air^88–90^, indoor air was identified as another hotspot, particularly for its role as a transport mechanism for resistant organisms. Unlike in sink drains, where water potentially supports bacterial metabolism and growth, bacteria in dry environments (e.g., doorsills) are more likely dormant^80^. Thus, we hypothesize that these bacteria are transferred to such environments and remain in the same state as when they were on human bodies. For example, bacteria detected on doorsills, including *A. radioresistens*, *S. maltophilia*, and *E. hormaechei*, may be carried on air currents and settle there along with biological debris (e.g., skin squames) shed by humans. Previous studies have implicated this phenomenon for *S. aureus*^91,92^. It is possible that settled bacteria could be re-suspended into the air and potentially infect patients. Although high-touch surfaces yielded fewer CHG-tolerant isolates compared to sink drains and air, they still hold value for monitoring due to frequent interactions. Note that the roles of environmental locations are dynamic within this interaction model, shifting as conditions and time change. For example, sink drains act as reservoirs when they receive inputs from hands, incoming water, and air, hosting microbes in biofilms. They can become sources of exposure when microbes are re-aerosolized^61^, or serve as transport mediums when contaminants flow through them to wastewater treatment plants. Similarly, the two hypotheses are also dynamic and condition-dependent.

Detection of CHG-tolerant, ARG-carrying, and/or opportunistic bacteria in environments does not necessarily translate to elevated risk of infection and antibiotic resistance due to several challenges and knowledge gaps^93,94^. First, more studies are needed to clarify the relationship between carrying ARGs and manifesting resistance phenotypes or increased tolerance. For instance, while *qacEdelta1* was a shared ARG in some high-CHG-MIC isolates (Fig. 4b-c), its presence was not exclusive to these, as it was also identified in two isolates with relatively low MICs (*P. aeruginosa*: 32 µg/mL, below its epidemiological cut-off value 50 µg/mL^12^; genus *Citrobacter*: ≤ 4 µg/mL; Table S4). In contrast, carrying *norA* (the other known CHG ARG) in *Staphylococcus epidermidis* appeared to associate with reduced susceptibility^25^. The *norA*-carrying isolate from our cohort showed an MIC of 4 µg/mL. Compared with previous reports^95^, this was 4.3 times higher than that observed in 33 blood isolates from 1965-1966, 2.1 times that of 290 more recent clinical isolates (post-2002), and comparable to that in four isolates from infected joint replacements. Future studies should aim to extend these qualitative phenomena observed in a limited sample size to quantitative analyses.

In addition to these ARGs previously reported to associate with CHG tolerance, we demonstrated from various angles that novel resistance mechanisms await discovery, and identified ARG and plasmid candidates (i.e., type-1 and type-3 *qacEdelta1*-carrying multidrug-resistant plasmids). Despite the richness of available data resources, we were challenged by the poor representation of environmental bacteria in current databases. For example, when searching the BLAST database for our *qacEdelta1*-carrying plasmid scaffolds, we found two top hits for *P. aeruginosa* with 100% query coverage. However, the maximum query coverage was only 46% for *Citrobacter spp.* and 20% for *C. metallidurans*. Expanding these databases and performing *de novo* gene annotations as complementary approaches would be beneficial in the future. To further connect carriage of ARGs to resistance phenotypes, future investigations should interrogate these candidates using functional metagenomics^96^, knockouts or cloning, as well as assess cross-resistance to medically relevant antibiotics via susceptibility tests (e.g., MIC assays).

Second, the clinical importance of detecting biocide resistance phenotypes in laboratories (e.g., using MIC assays) is not sufficiently understood and lacks consensus. The clinical relevance includes both disinfection efficacy and the evolution of cross-resistance to antibiotics, particularly in opportunistic pathogens. Several terms - resistance, tolerance, and reduced susceptibility - are used ambiguously in the field to describe these phenotypes, but their definitions and applications are not well standardized. Moreover, there is ongoing debate over whether MIC is the appropriate metric for assessing biocide efficacy^97^. MIC, which was originally developed for antibiotics, measures the ability of a compound to inhibit bacterial growth over 16-24 hours, whereas biocides are often applied at high concentrations for brief periods during skin antisepsis or environmental disinfection. Special cases exist, such as CHG bathing, where we seek both immediate antiseptic effects and longer-term benefits from residual CHG on the skin for up to 24 hours. To address some of these ambiguities, studies have proposed new frameworks^98^ and metrics (e.g., the minimum duration for killing) to complement MIC. However, these alternatives lack sufficient benchmarking and evaluation of generalizability. As surveillance expands, generalizability becomes increasingly important - a limitation the current gold standard, MIC, also faces. For example, clinical breakpoints for chlorhexidine, as with many other antimicrobials, have not yet been established^27^. Reaching consensus on clinical breakpoints and even epidemiological cut-offs is challenging. For instance, we used 18.75 µg/mL as the cut-off for CHG tolerance, as it was indicated as a threshold CHG skin concentration^3^; however, a recent study has shown disagreement^99^. Nonetheless, CHG concentrations used in clinical applications on skin and mucosal surfaces are much higher than the highest MICs identified in our environmental isolates.

Large-scale environmental surveillance introduces additional challenges. During MIC assays, despite following the CLSI standards^100^ as closely as possible, we could not confirm that all inocula were standardized to 10^4^ CFU per 5- to 8-mm diameter spot (Methods) because the taxonomy and growth characteristics of isolates were unknown at the time of testing. Furthermore, it remains unclear whether the standard CFU threshold has the same clinical relevance for environmental isolates as it does for clinical isolates, which were used to develop the standard. The limitations of MIC extend beyond the lab. In environmental settings, microbes rarely exist in isolation or as free-living cells; rather, they often form biofilms within complex communities where synergistic/competitive interactions and physical protection may occur. As a result, assays that simulate their in-situ states may offer more accurate insights. Alternative approaches, such as examining contact times necessary to achieve a reduction threshold^97^ or using microcosm experiments, like those employed in our study, may provide better approximations of real-world conditions. For instance, we examined bacterial survival following 24-h CHG exposure on surface coupons (Fig. 1c), which better approximates in-situ conditions than liquid media assays. However, this approach requires optimization. To ensure technical feasibility, we used a higher starting bacterial density than is typically found on uncontaminated hospital surfaces. This version simulates a scenario where residual CHG is present on a surface, and a high burden of pathogens is introduced (e.g., through sneezing). More research is needed to develop consensus methods that are applicable to real-world scenarios.

Furthermore, researchers should be more explicit when communicating findings. For example, while chlorhexidine digluconate is the most used formula, other forms, such as chlorhexidine acetate, exist. We have encountered instances where the type of formula was not explicitly reported in the literature, hindering normalized evaluations through reviews and meta-analyses^101^.

Despite these challenges, we conducted surveillance of CHG tolerance in built-environment isolates, a practice less common than in clinical and animal isolates. The MICs of environmental isolates are generally considered lower (Table S6)^16,17,19,22–24,102,103^. However, bacteria isolated in our study, especially those from sinks, exhibited CHG MICs at the high end of the reported spectrum^12,15,27,101,104–108^. Although further investigation is needed to determine whether our isolates represent a statistically significant increase in tolerance, the results suggest that the MICU environment harbors CHG-tolerant microbes, raising a potential warning sign.

Moving forward, we should further scrutinize the environmental hotspots (e.g., sink drains, air) and establish models to quantify the complex human-environment and chemical-microbe interplay. First, combining broad-scope studies with more focused, larger-sized studies is recommended. We should collect denser longitudinal samples and richer metadata; for example, a logical next step would be to survey chlorhexidine concentrations in MICUs. Second, profiling these samples using multimodal approaches, such as combining multi-omic sequencing of communities with whole-genome sequencing of isolates, could provide deeper insights. Third, pairing environmental and human microbiota could uncover more nuanced connections. Furthermore, as our understanding in MICUs advances, studies could be expanded to other buildings (e.g., entire hospitals, veterinary facilities, homes, athletic facilities) to determine whether findings surrounding CHG tolerance are unique to MICUs, hospitals, or built environments in general.

In summary, by combining microcosm experiments with field surveys, we provided a comprehensive depiction of CHG tolerance in MICU environments, connecting chemical, microbial, and molecular perspectives. Understanding these interactions could reveal potential intervention opportunities for infection prevention and lead to actionable recommendations for regulations and healthcare policy. Given the critical value of CHG in healthcare and the dearth of viable alternatives, proactive measures are important, such as integrating environmental management with clinical interventions, expanding stewardship programs, and exploring alternatives to CHG. Furthermore, due to chlorhexidine’s versatile applications, the insights gained from this study could benefit a broader audience, informing stewardship practices in veterinary hospitals and other built environments, as well as guiding consumer use of antiseptics and disinfectants.

## Methods

### Chlorhexidine persistence on surfaces

We investigated the chlorhexidine persistence on three surface materials commonly found in hospital (laminate wood, plastic [high-density polyethylene; HDPE], metal [stainless steel]) in response to six different cleaning practices used commonly on inanimate hospital surfaces (no cleaning, clean with water, disinfect with ethanol, bleach, peracetic acid, or benzalkonium chloride) (Fig. S1).

#### Microcosm setup

Each surface was cut into 4”×8” coupons, and each coupon was divided into 1”×1” sections by drawing markers. The coupons were UV sterilized for 15 minutes before applying 2% chlorhexidine digluconate (CHG, IUPAC name: (1E)-2-[6-[[amino-[(E)-[amino-(4-chloroanilino)methylidene]amino]methylidene]amino]hexyl]-1-[amino-(4-chloroanilino)methylidene]guanidine;(2R,3S,4R,5R)-2,3,4,5,6-pentahydroxyhexanoic acid) solution (w/v, Sigma-Aldrich, St. Louis, MO, USA). The concentration 2% was selected to represent commercial CHG wipes used in patient bathing^109,110^. We first folded a Kimwipe (Kimberly-Clark, Irving, TX, USA) in half three times to get a square about 2”×2” and 8 layers thick. We then pipetted 1 mL CHG onto the Kimwipe and wiped in a zigzag pattern along the long axis of the surface, such that the whole surface was covered after 5 passes. Wiping was repeated with a second Kimwipe from the opposite direction. Once the surface dried, an initial CHG measurement was taken in biological triplicate, representing 0 min. Each 1”×1” section was swabbed using a nonsterile cotton swab (Puritan, Guilford, ME, USA) premoistened in 0.8 mL deionized water. The swabbing was conducted with consistent pressure and speed in two directions (up and down, left and right) for 15 s total. The swab was then stored in the deionized water for extraction and CHG concentration measurement.

Simulated cleaning practices were applied to CHG-treated surfaces using the same wiping technique. A group of surfaces that were not wiped served as baseline controls. The wiping solutions included deionized water, 70% ethanol (Decon Laboratories, King of Prussia, PA, USA), 10% bleach (Clorox, Oakland, CA, USA), 0.2% peracetic acid (Sigma-Aldrich, St. Louis, MO, USA), and 0.26% benzalkonium chloride (MP Biomedicals, Santa Ana, CA, USA; C8-18)^111^. The CHG concentrations were measured in biological triplicate at 5 additional timepoints (10 min, 1 h, 3 h, 6 h, and 24 h) following the same method. We selected 24 h as an experimental cycle because in MICU, patients were bathed with CHG wipes on a daily basis. The coupons were kept in light-proof compartments when not being wiped or swabbed. This was repeated for each disinfectant and each surface for 8 trials, yielding 24 biological replicates in total.

#### Swab extraction and CHG concentration measurement

CHG on swabs was extracted into water following an adaptation of CDC’s Swab Extraction Method^94,112,113^. The Falcon tube was vortexed at maximum speed for 1 min and shaken at 180 rpm and 25 °C for 10 min, followed by careful squeezing against the tube wall and removal of the swab. CHG concentration was determined by a previously reported colorimetric assay with modifications^3,114,115^. We mixed 500 μL CHG solution with 500 μL hexadecyltrimethylammonium bromide (0.01 g/mL in deionized water, Sigma-Aldrich, St. Louis, MO, USA) and 200 μL sodium hypobromite (Aqua Solutions, Deer Park, TX, USA). Absorbance of the mixture was measured at 262 nm using a spectrophotometer (UV-2450, Shimadzu, Kyoto, Japan).

#### Quantification of CHG recovery rates from surfaces and application efficiency to surfaces

We examined the CHG recovery rates from different surface materials using the same swabbing technique and accounted for these rates in calculating the CHG concentration on surfaces (Table S7). After UV sterilization, we pipetted 100 μL 2% CHG onto a 1”×1” square for a total of 10 replicates each surface type. Additionally, 100 μL 2% CHG was directly pipetted onto a swab to investigate the recovery rate of the swab extraction. When we directly applied CHG onto swabs, 85.46% was recovered in the measurement. For the tested surfaces, metal had a comparably high recovery rate (83.04%), indicating that almost all the CHG (97.16%) on a metal surface can be collected by swabbing. In contrast, plastic and laminate wood both retained CHG in sampling, with recovery rates of 69.79% and 55.58%, respectively.

CHG transfer rates from the Kimwipe to the surfaces in application was quantified afterwards (Table S8). Upon wiping a surface with a CHG wipe, we found that only 4-7% CHG on the wipe was transferred onto the surfaces. In comparison to the recovery rates from surfaces by swab sampling, the transfer rates showed an opposite trend across surface materials. While metal had the highest recovery rate in sampling, it captured the least CHG in application. Furthermore, the values were within the range of disinfectants released from ready-to-use Towelettes^116^, validating that our application technique mimics the usage of commercial disinfectants.

### Viability assay of bacterial isolates on CHG-contaminated surfaces

We tested the survival of bacterial isolates on plastic surfaces after exposure to various concentrations of CHG for 24 h in biological triplicate. Four bacterial isolates (*Escherichia coli* [ATCC 25922], *Klebsiella pneumoniae* [ATCC 13883], *Klebsiella variicola* [20-20012]^37^, and *Staphylococcus aureus* [ATCC 29213]) were selected as model strains. Plastic was selected as a representative surface because it is a common material of equipment in the immediate vicinity of patients and healthcare workers (e.g., bedrail, nurse call button, keyboard, mouse). Five different CHG concentrations (31.00, 15.50, 3.10, 0.31, 0 [positive control] μg/cm^2^) were tested to cover the range of CHG persisting on surfaces under disinfection and cleaning.

To prepare bacterial inocula, the second passage of the stock culture were grown at 37°C to the late exponential phase and harvested at 3000 rpm for 1 min. The pellet was then resuspended in phosphate-buffered saline (PBS). The bacterial cell density in the inocula was determined by spread plating. To prepare surface coupons, HDPE plastic was cut into 2”×2” squares, UV sterilized for 15 min, and placed in a petri dish. CHG was applied to the center 1”×1” of the coupon by pipetting 100 μL solution and spreading evenly with the pipette tip. Subsequently, 100 μL isolate suspension was applied using the same technique^117^. An additional coupon inoculated with 100 μL PBS was included as the negative control. Surfaces were kept in dark at room temperature.

After 24 h, bacteria were sampled from surfaces and the surviving numbers were counted. Each surface was first dry swabbed for 5 s and wet swabbed twice for 7.5 s each time after rinsing in 1 mL PBS. Finally, the swab was squeezed against the tube wall to expel any residual liquid^117^. Samples were diluted and 100 μL of each dilution was spread plated onto TSA plates. The plates were incubated at 37°C for 24 hours to count the colonies.

#### Field survey in a Medical Intensive Care Unit

##### Sample collection

We collected 219 swab samples from seven different locations (e.g., bedrails, nurse call buttons, doorsills, keyboards, light switches, proximal sink drains; Table S3) in the MICU at Rush University Medical Center over two sampling events (February and July, 2018) (Fig. S2), including negative field controls and negative media controls. The MICU was selected because intensive care units are known to have high prevalence of antibiotic-resistant organism colonization and infection, and because patients in the MICU received daily skin cleansing with 2% CHG-impregnated cloths (Sage 2% Chlorhexidine Gluconate Cloths, Stryker, Portage, MI, USA)^118,119^, which gives this environment a higher probability but also a lower threshold for acceptable risk of containing antibiotic-resistant and CHG-tolerant organisms. Sampling locations were selected to capture a variety of surfaces reflecting different ways in which people interact with the surface^120^ and time-integrated aggregates of what was in the air (i.e., doorsills)^121^. Sites were further selected to facilitate comparison with other hospital surveillance studies^122^ and to keep a relatively balanced number of samples from each touch frequency group. Touch frequency categorization included bedrails and nurse call buttons as high-touch, keyboards as medium-touch, light switches as low-touch surfaces, and doorsills as no-touch. Sink drains were labeled as themselves. Touch frequency designations were based on interviews with healthcare professionals and prior evidence from the literature^123–126^.

Swabbing was conducted following CDC’s Environmental Surface Sampling Swab Contact Method. Briefly, we used three sterile Nylon Flocked Dry Swabs (COPAN Diagnostics, Murrieta, CA, USA) premoistened with PBS to increase the recovery of biomass. Surfaces were swabbed in their entirety three times with consistent pressure and speed. Each time, we rotated the swabs and switched their order. Swabs were stored in 15 mL tubes with PBS and transferred to the lab on ice within 24 hours for immediate processing. Temperature, relative humidity, isolation level, and patient mobility for each patient room were recorded. The surface area of each sampling location was measured (Table S3).

##### Swab extraction, sample cultivation, and initial screening

Biomass on swabs was extracted into PBS following an adaptation of CDC’s Swab Extraction Method^94,112,113^. Briefly, the Falcon tubes with swabs inside were vortexed at maximum speed for 10 s, shaken at 180 rpm and 25 °C for 10 min, and then vortexed again for 10 s, followed by careful squeezing against the side of the tube and removal of the swabs.

The samples were then subjected to cultivation and screening following a protocol modified from a previous study^127^ (Fig. S3). Inocula were diluted to obtain between 30 and 300 colonies per plate. 100 µL of inoculum was spread plated on tryptic soy agar supplemented with 4 mg/L itraconazole (TSAI) and incubated at 25°C for 4 days. When observable growth was detected, morphologies of colonies were characterized using standardized ontologies for streamlined analysis, and a representative number of colonies belonging to each morphology type were picked and stored in glycerol at -80°C. These plates were then replica plated onto blood agar and chlorhexidine agar plates for initial screening^128^. Chlorhexidine agar plates contained 10.56 µg/mL chlorhexidine (equivalent to 18.75 µg/mL chlorhexidine digluconate, Sigma-Aldrich, St. Louis, MO, USA). *Staphylococcus aureus* ATCC 25923, *Staphylococcus epidermidis* ATCC 12228, and *Pseudomonas putida* 56A10 (in-house ID)^129^ were used as control strains in this chlorhexidine screening process. The chlorhexidine concentration was selected for initial tolerance screening because it was reported as the minimum effective concentration associated with significantly decreased colony counts of Gram-positive bacteria on the skin of patients who were bathed daily with CHG in the MICU. In addition, all the patient skin isolates recovered in the study had MICs below 18.75 µg/mL CHG^3^.

##### Antimicrobial susceptibility testing

We confirmed and refined chlorhexidine tolerance observed in the initial screen using a simplified agar dilution method adapted from the CLSI standard M07Ed11^100^. Briefly, agar dilution plates with different concentrations of CHG (0, 4, 8, 16, 32, 64, 128, 256, 512 μg/mL) were made with Mueller-Hinton Agar (MHA; Sigma-Aldrich, St. Louis, MO, USA) and chlorhexidine digluconate solution (20% w/v, Sigma-Aldrich, St. Louis, MO, USA). Each isolate was first inoculated from frozen stock in 100 µL TSB in a sterile 96-well clear round-bottom not-treated microplate (Corning, NY, USA) and incubated at 37 °C for 24 h, yielding F1. F1 was subcultured in 100 µL TSB with a 96-pin microplate replicator (transfers 1 µL inoculum each pin, Boekel Scientific, Feasterville-Trevose, PA, USA) and incubated for another 24 h, yielding F2. F2 was inoculated onto surfaces of the agar dilution plates in biological triplicate with the 96-pin replicator, starting from the lowest concentration to the highest. MICs were determined after incubating the plates at 37 °C for 20 h. Negative controls and two quality control (QC) strains were included in every batch. The two QC strains were both MRSA with CHG MICs 1-2 and 8 µg/mL, respectively^37^.

### Genomic exploration of isolates recovered in the MICU

#### Whole-genome sequencing

A subset of 63 isolates were selected for whole-genome sequencing in two batches. Three negative controls were processed in parallel to samples throughout the entire pipeline, and all isolates were sequenced at 2×150 bp. The first batch was conducted by the Broad Institute using a fluorescent dye-based method on an Illumina HiSeq platform as previously described^129^. The second batch was performed at the NUSeq Core Facility at Northwestern University on an Illumina NovaSeq 6000 platform using an SP flow cell, and the sequencing library was prepared using the Illumina Nextera XT DNA Sample Preparation Kit following the kit protocol. The average coverage depth was 214.9× (87.5× ∼ 536.5×) by estimation assuming 5 Mb for bacterial genome sizes.

#### Genome assembly, taxonomic identification, and phylogenetic analysis

The analysis pipeline was illustrated in Fig. S8. Raw reads were first quality checked by FastQC^130^ (v0.11.5) and cleaned by fastp^131^ (v0.23.2). Genomes were assembled using SPAdes^132^ (v3.15.0) with the assembly quality assessed by CheckM^133^ (v1.2.2) and QUAST^134^ (v5.2.0). In the genome assembly, SPAdes parameters --careful and --isolate were compared and selected according to the quality of the resulted scaffolds. Integrating insights from the research community^135^, --careful was used for all samples except isolate S33. We then performed taxonomic assignments using KmerFinder^136–138^ (v3.0.2) with the database version 20211017 and PhyloPhlAn^139^ (v3.0.36) metagenomic SGBs with the database SGB.Jul20. Isolates with inconsistent assignments were further examined using autoMLST^140^ *de novo* mode and MiGA TypeMat^141^. Phylogenetic trees among bacteria isolates were constructed with PhyloPhlAn (v3.0.36) on the low diversity setting using the assembled scaffolds. The strain-level relationship between isolates was determined using fastANI for ANI values ≥ 99.99%^59,60^. The phylogenetic tree among plasmid scaffolds (Fig. 6c) was constructed with Clustal Omega^142^.

#### Analysis of antimicrobial resistance genes

ARGs were predicted using Resistance Gene Identifier (RGI, v6.0.2) coupled with the Comprehensive Antibiotic Resistance Database (CARD, v3.2.7)^76^. Only ARGs under perfect and strict paradigms were included in the downstream analyses. ARGs presented in this study were named according to the curated CARD short names (i.e., Best_Hit_ARO output by RGI)^76^. Briefly, if the original gene name is less than 15 characters, the CARD short name is identical; otherwise, it has been abbreviated by CARD curators specifically to identify the proper gene or protein name. Mapping between a unique CARD short name and the corresponding metadata can be found in the CARD ontology database (https://card.mcmaster.ca/), including AMR gene families, drug classes, resistance mechanisms, resistomes, and homolog sequences. Mobile genetic elements were predicted using geNomad^143^ (v1.5.2), MobileElementFinder^144^ (mge_finder v1.1.2, mgedb v1.1.1, blastn v2.14.0+), and VRprofile2^145^ with default parameters. In MobileElementFinder outputs, putative composite transposons were excluded to avoid false-positive predictions. To infer the mobility of ARGs, they were classified as MGE-carried, MGE-associated, or MGE-unassociated according to their relative location to MGEs. Specifically, ARGs located within MGEs were classified as MGE-carried, whereas those situated within a 5 kb proximity to an MGE were considered MGE-associated. Both MGE-carried and MGE-associated ARGs were deemed mobile^68,146^. To prioritize ARGs, health risk indices from Zhang et al. were employed, which they crafted by integrating factors of human accessibility, mobility, pathogenicity and clinical availability^68^. To contextualize our findings within the framework of known CHG resistance mechanisms, we carried out a targeted review and curated a list of CHG ARGs, annotating each with a confidence level based on whether the evidence was correlational or causal (Table S9).

### Statistical analysis

Statistical analyses and data visualization were conducted in R (v4.0.4)^147^ with packages such as dplyr^148^, stringr^149^, rstatix^150^, ggplot2^151^, ComplexHeatmap^152^, and gggenes^153^. Differences between groups were determined by Student’s *T*-test with p-values adjusted by the Benjamini-Hochberg method or ANOVA coupled with Tukey’s post hoc test. p ≤ 0.05 was defined as statistically significant. Significance codes are as follows: p > 0.05 (ns), 0.01 < p ≤ 0.05 (*), 0.001 < p ≤ 0.01 (**), 0.0001 < p ≤ 0.001 (***), and p ≤ 0.0001 (****).

## Data Availability

The raw whole genome sequencing data of bacteria isolates are available in the NCBI SRA repository under BioProject number PRJNA1169385. The data will be accessible upon manuscript acceptance. Source code for the analyses are available under MIT license at https://github.com/hartmann-lab/Chlorhexidine_Tolerance_Hospital_Environments. Bacteria isolates are available upon request.

## Supplementary figures

**Fig. S1 Coupons used in the microcosm experiments examining chlorhexidine persistence.** Grids on the coupons indicate specific swabbing sites.

**Fig. S2 a) Floor plan of the Medical Intensive Care Unit at the sampled hospital. b) Illustrations of the swabbed area for the sampled environmental locations.**

**Fig. S3 Cultivation and screening pipeline.** TSAI is tryptic soy agar (TSA) supplemented with 4 mg/L itraconazole. TSA+CHX is TSA containing 10.56 µg/mL chlorhexidine powder (equivalent to 18.75 µg/mL CHG).

**Fig. S4 Absolute bioburden of bacteria (log_10_CFU) in the entirety of the dry surfaces and by swabbing the proximal sink drain.** The bacteria CFUs were not normalized by sampling area. Differences between groups were determined by unpaired *t*-tests with the Benjamini-Hochberg adjustment. To avoid mis-interpreting data with small colony counts, we empirically set the limit of quantification (LOQ) of our culture efforts as colony counts = 3. Values below this threshold were recorded as half of LOQ (i.e., 1.5). Nurse call button is abbreviated as nurse call in the figure.

**Fig. S5 No significant differences were observed in: a)** Unit area bioburden of bacteria between the two sampling seasons, **b)** Unit area bioburden of bacteria between patient rooms with contact isolation and those without, and **c)** Proportions of CHG-tolerant bacteria (tolerating 18.75 µg/mL CHG at 25°C) between contact isolation levels of patient rooms. Cultivation temperature was 25°C for all cases. Nurse call button is abbreviated as nurse call in the figure.

**Fig. S6 A phylogenetic tree of all sequenced isolates.**

**Fig. S7 Potential dissemination of opportunistic pathogens across multiple MICU rooms.** Patient rooms were deidentified. Each panel shows one species with multiple isolates detected in sink drains of different MICU rooms during the same sampling event.

**Fig. S8 Analysis pipeline of whole-genome sequences.**

## Supplementary tables

**Table S1 Epidemiological cut-off values for CHG resistance.**

**Table S2 Statistical testing results of CHG persistence on surfaces.** List of abbreviations for cleaning: C="No liquid", W= "Water", Q= "Benzalkonium chloride", P= "Peracetic acid", B= "Bleach", E= "Ethanol"

**Table S3 Sample collection metadata. a)** Specific sampling locations within the MICU **b)** Distribution of Number of collected samples for each sampling event **c)** List of Abbreviations for sampling locations and space types **d)** Surface area of sampling locations

**Table S4 Antibiotic resistance genes (ARGs, curated CARD short name) predicted using Resistance Gene Identifier (RGI) and Comprehensive Antibiotic Resistance Database (CARD) under Perfect and Strict paradigms**, with further annotations of ARG health risk (blue), chlorhexidine MICs (green), taxonomic assignments (orange), sample collection and cultivation results (purple), mobile ARGs (gray). List of abbreviations for Space Type: MD = “Medication room”, PT = “Patient room”, CM = “Communicating space”, ST = “Staff-only restroom.” Room numbers were deidentified by randomly generating unique numbers.

**Table S5 Additional information of *qacEdelta1*-carrying multidrug-resistant plasmid scaffolds. a)** Metadata of *qacEdelta1*-carrying plasmid scaffolds. **b)** Intra-species similarity of *qacEdelta1*-carrying plasmid scaffolds determined by blastn. **c)** Inter-species similarity of *qacEdelta1*-carrying plasmid scaffolds determined by blastn. **d)** Assessment of the completeness of plasmid scaffolds. **e)** The top BLAST hits for type-1 plasmid scaffolds matched two plasmids from clinical *P. aeruginosa* strains - one from a bloodstream infection and the other from sputum - both collected in Chicago.

**Table S6 Reports of the highest CHG MICs (µg/mL) for environmental isolates.** Bacteria isolated in our study, especially those from sinks, exhibited MICs at the high end of this spectrum.

**Table S7 Recovery rate of CHG on different surface materials.** The recovery rate measured for surfaces was the integrative rate, representing the overall impacts of swabbing on surfaces, extracting CHG from the swab, and all the loss in the process.

**Table S8 Transfer rate of chlorhexidine digluconate (CHG) from Kimwipe to surfaces by wiping.**

**Table S9 A targeted review of antibiotic resistance genes (ARGs) conferring resistance to chlorhexidine.** CHX resistance: Y, confer resistance to CHX; N, do not confer resistance; C, controversial with conflicting results. Correlation & causality: Y, evidence available; N, evidence not available; NA, not applicable.

## Acknowledgements

This study was partially supported by the Searle Leadership Fund and the Northwestern University 2020 NUSeq Pilot Project Program. Additionally, the computational resources and staff contributions provided by the Quest high performance computing facility at Northwestern University, which is jointly supported by the Office of the Provost, the Office for Research, and Northwestern University Information Technology, were instrumental.

JS was supported by Northwestern University Terminal Year Fellowship from the Richter Memorial Fund. Undergraduate researchers received support from Northwestern University Undergraduate Research Grants: AW from the Buffett Institute, MK from the Weinberg College of Arts and Sciences Summer Undergraduate Research Grant, and YW and YS from the Office of Undergraduate Research Summer Undergraduate Research Grant.

We are deeply thankful to Teppei Shimasaki, Khaled Aboushaala, Thelma Dangana, Yoona Rhee and Alexander G. McFarland for their help in sample collection and coordination. Our appreciation extends to the clinical staff of the Medical Intensive Care Unit at Rush University Medical Center. We gratefully acknowledge Adam J. Glawe and James Lindsay for their contributions to microbial cultivation. Finally, we would like to thank the anonymous reviewers whose insights significantly improved this manuscript.

