## Supplementary material for "Hospital environments harbor chlorhexidine tolerant bacteria potentially linked to chlorhexidine persistence in the environment": Fig. S

### Supplementary figures S1-S8

### Hospital environments harbor chlorhexidine tolerant bacteria potentially linked to chlorhexidine persistence in the environment

Jiaxian Shen[1], Yuhan Weng[1], Tyler Shimada[1], Meghana Karan[1], Andrew Watson[1], Rachel L. Medernach[2], Vincent B. Young[3], Mary K. Hayden[2], Erica M. Hartmann[1,4,5,*]

[1]Department of Civil and Environmental Engineering, McCormick School of Engineering, Northwestern University, USA

[2]Division of Infectious Diseases, Department of Internal Medicine, Rush University Medical Center, USA

[3]Department of Internal Medicine/Division of Infectious Diseases, Department of Microbiology & Immunology, University of Michigan Medical School, USA

[4]Center for Synthetic Biology, Northwestern University, USA

[5]Department of Medicine/Division of Pulmonary Medicine, Feinberg School of Medicine, Northwestern University, USA

Supplementary information includes 8 figures and 9 tables.

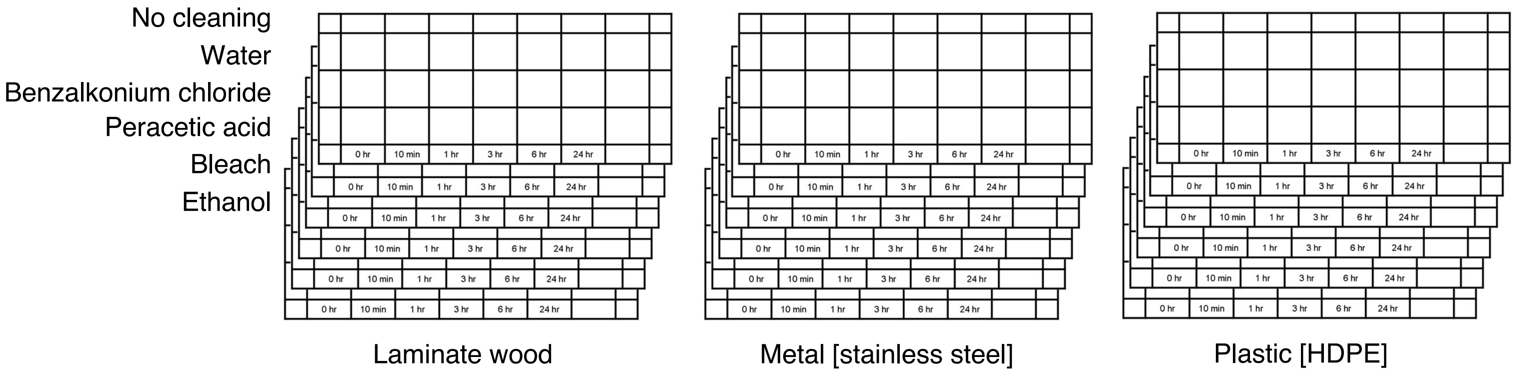


**Fig. S1 Coupons used in the microcosm experiments examining chlorhexidine persistence.** Grids on the coupons indicate specific swabbing sites.

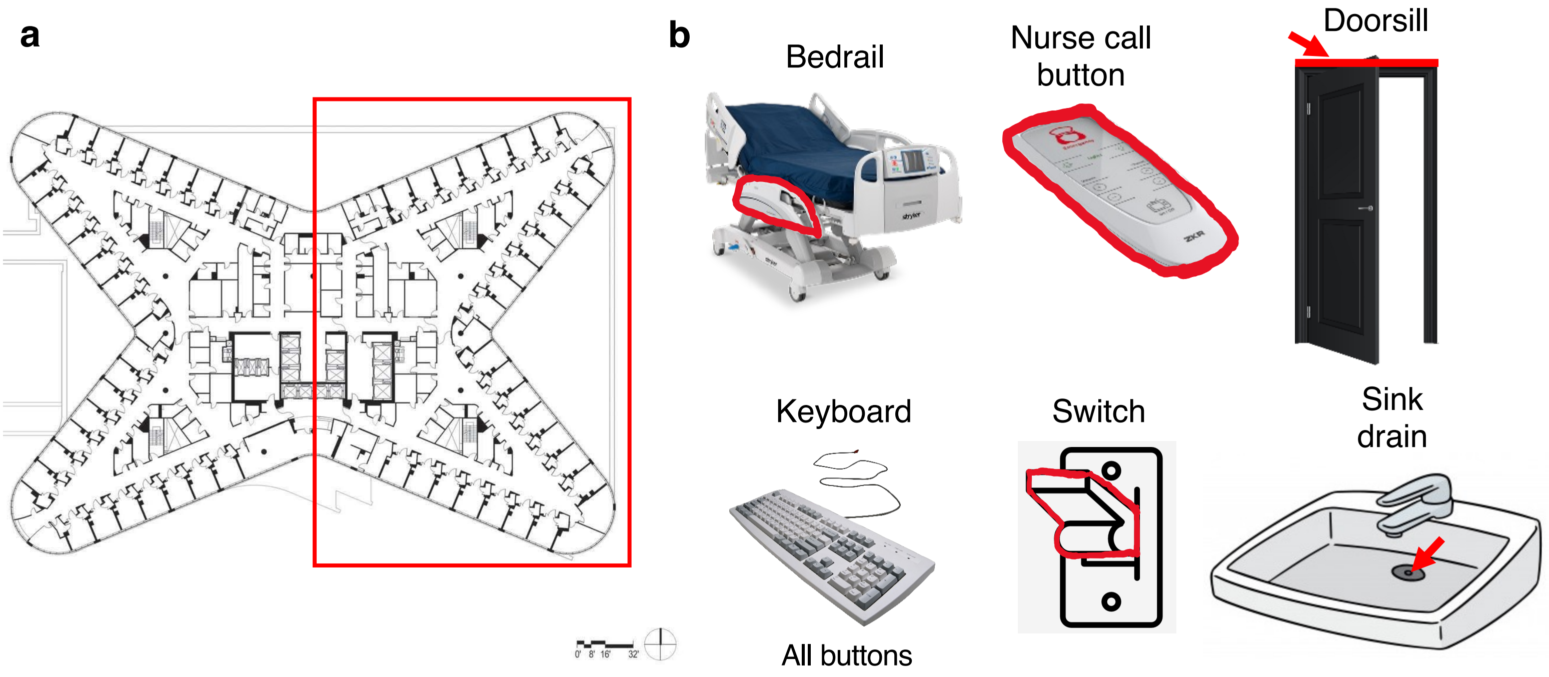


**Fig. S2 a) Floor plan of the Medical Intensive Care Unit at the sampled hospital. b) Illustrations of the swabbed area for the sampled environmental locations.**

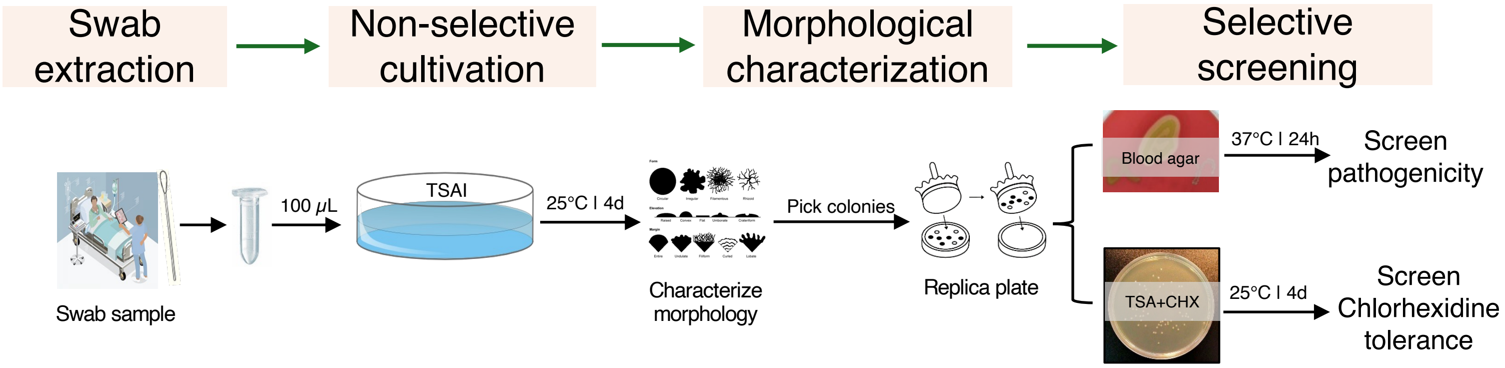


**Fig. S3 Cultivation and screening pipeline.** TSAI is tryptic soy agar (TSA) supplemented with 4 mg/L itraconazole. TSA+CHX is TSA containing 10.56 µg/mL chlorhexidine powder (equivalent to 18.75 µg/mL CHG).

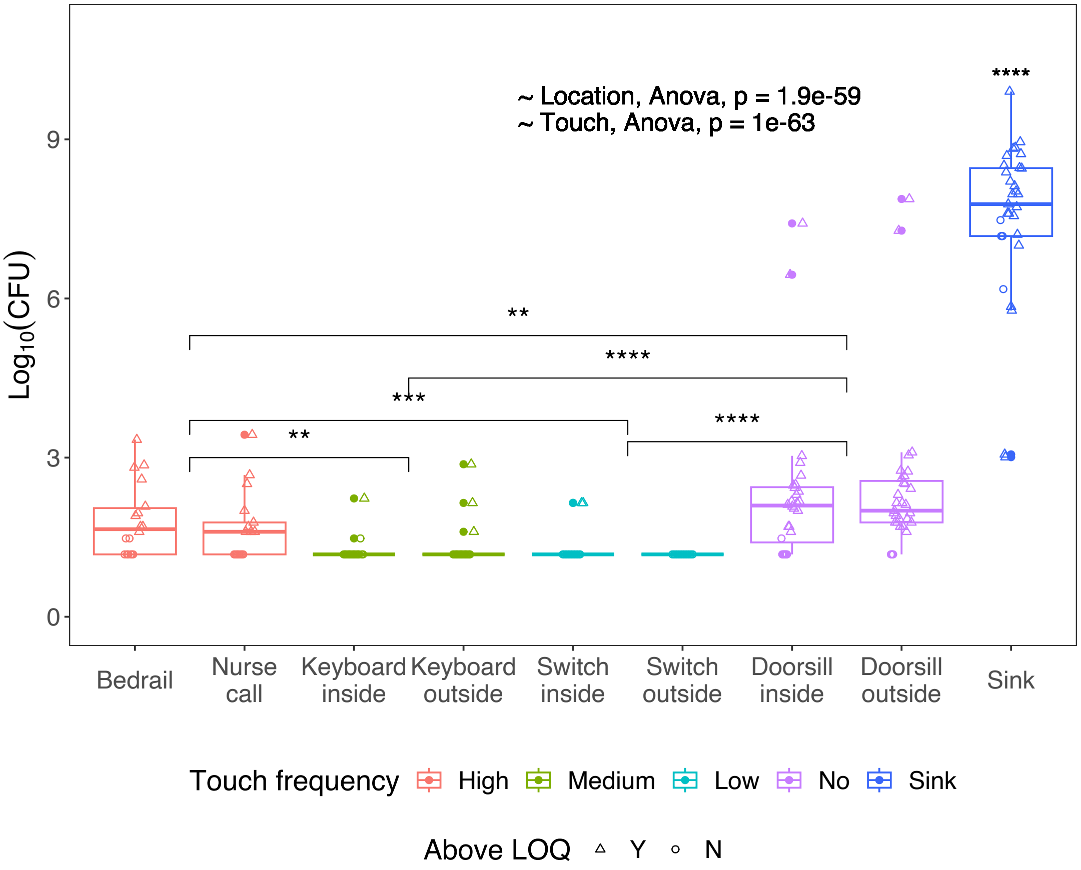


**Fig. S4 Absolute bioburden of bacteria ($\log_{10}$CFU) in the entirety of the dry surfaces and by swabbing the proximal sink drain.** The bacteria CFUs were not normalized by sampling area. Differences between groups were determined by unpaired *t*-tests with the Benjamini-Hochberg adjustment. To avoid mis-interpreting data with small colony counts, we empirically set the limit of quantification (LOQ) of our culture efforts as colony counts = 3. Values below this threshold were recorded as half of LOQ (i.e., 1.5). Nurse call button is abbreviated as nurse call in the figure.

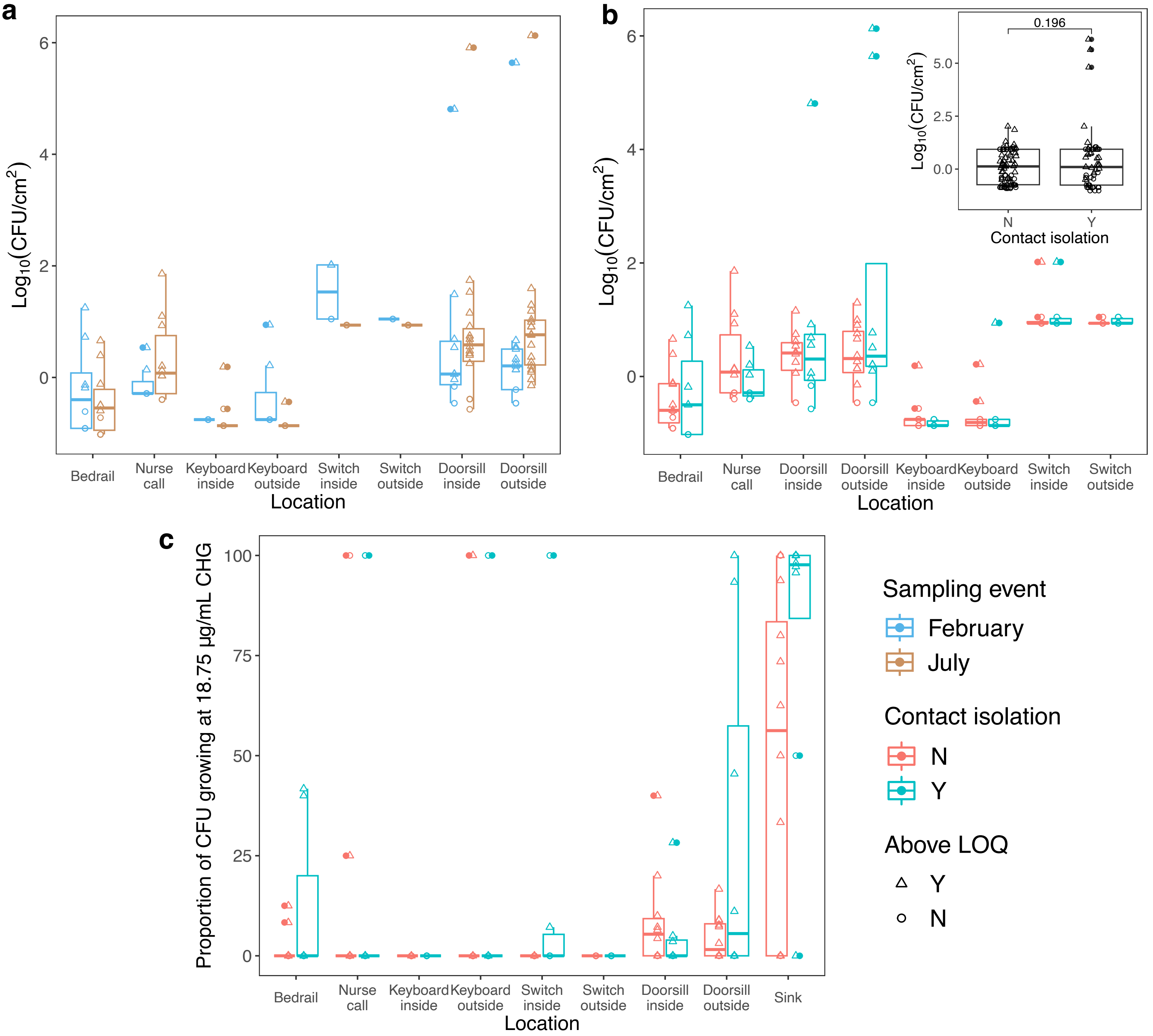


**Fig. S5 No significant differences were observed in: a)** Unit area bioburden of bacteria between the two sampling seasons, **b)** Unit area bioburden of bacteria between patient rooms with contact isolation and those without, and **c)** Proportions of CHG-tolerant bacteria (tolerating 18.75 µg/mL CHG at 25°C) between contact isolation levels of patient rooms. Cultivation temperature was 25°C for all cases. Nurse call button is abbreviated as nurse call in the figure.

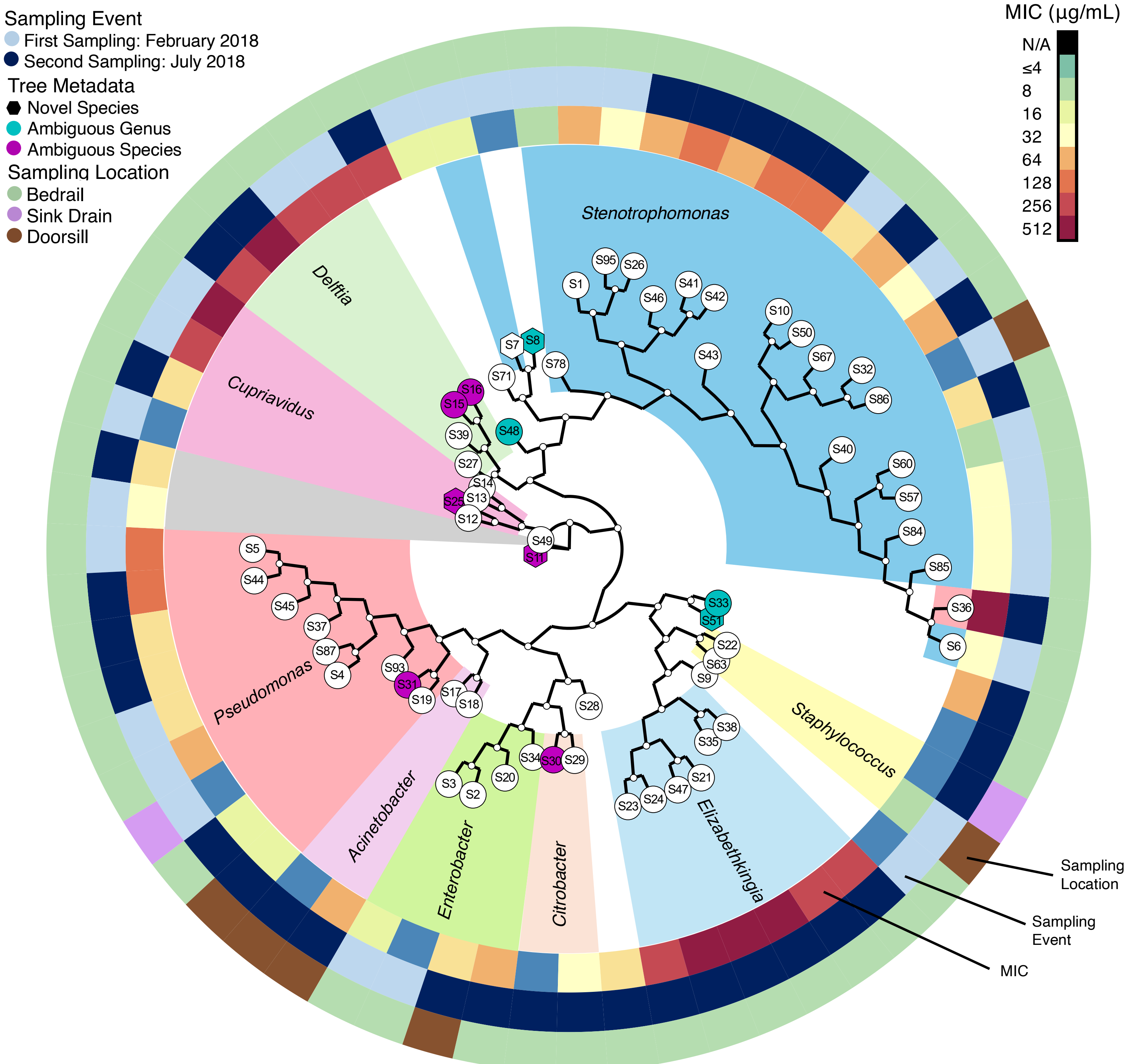


S49 *Achromobacter aegrifaciens*
S17 *Acinetobacter radioresistens*
S18 *Acinetobacter radioresistens*
S30 *Citrobacter amalonaticus or Citrobacter freundii*
S29 *Citrobacter freundii*
S13 *Cupriavidus metallidurans*
S14 *Cupriavidus metallidurans*
S12 *Cupriavidus pauculus*
S27 *Delftia tsuruhatensis*
S39 *Delftia tsuruhatensis*
S15 *Delftia tsuruhatensis or Delftia acidovorans*
S16 *Delftia tsuruhatensis or Delftia acidovorans*
S48 *Delftia tsuruhatensis, Delftia acidovorans or Stenotrophomonas maltophilia*
S21 *Elizabethkingia miricola*
S23 *Elizabethkingia miricola*
S24 *Elizabethkingia miricola*
S35 *Elizabethkingia miricola*
S36 *Elizabethkingia miricola*
S38 *Elizabethkingia miricola*
S47 *Elizabethkingia miricola*
S34 *Enterobacter asburiae*
S20: *Enterobacter hormaechei*
S2: *Enterobacter kobei*
S3: *Enterobacter kobei*
S28: *Klebsiella michiganensis*
S33: *Ochrobactrum anthropi or Brucella anthropi*
S4: *Pseudomonas aeruginosa*
S5: *Pseudomonas aeruginosa*
S37: *Pseudomonas aeruginosa*
S44: *Pseudomonas aeruginosa*
S45: *Pseudomonas aeruginosa*
S87: *Pseudomonas aeruginosa*
S93: *Pseudomonas azotifigens*
S19: *Pseudomonas putida*
S71: *Pseudomonas stutzeri*
S9: *Sphingobacterium siyangense*
S22: *Staphylococcus debuckii*
S63: *Staphylococcus epidermidis*
S1: *Stenotrophomonas maltophilia*
S6: *Stenotrophomonas maltophilia*
S10: *Stenotrophomonas maltophilia*
S26: *Stenotrophomonas maltophilia*
S32: *Stenotrophomonas maltophilia*
S40: *Stenotrophomonas maltophilia*
S41: *Stenotrophomonas maltophilia*
S42: *Stenotrophomonas maltophilia*
S43: *Stenotrophomonas maltophilia*
S46: *Stenotrophomonas maltophilia*
S50: *Stenotrophomonas maltophilia*
S57: *Stenotrophomonas maltophilia*
S60: *Stenotrophomonas maltophilia*
S67: *Stenotrophomonas maltophilia*
S78: *Stenotrophomonas maltophilia*
S84: *Stenotrophomonas maltophilia*
S85: *Stenotrophomonas maltophilia*
S86: *Stenotrophomonas maltophilia*
S95: *Stenotrophomonas maltophilia*
S51: *novel species, f_Rhizobiaceae, g_Pseudorhizobium or g_Rhizobium*
S8: *novel species, f_Xanthomonadaceae, g_Stenotrophomonas*
S11: *novel species, g_Achromobacter*
S25: *novel species, g_Cupriavidus*
S7: *novel species, g_Stenotrophomonas*
S31: *undetermined species, g_Pseudomonas*

**Fig. S6 A phylogenetic tree of all sequenced isolates.**

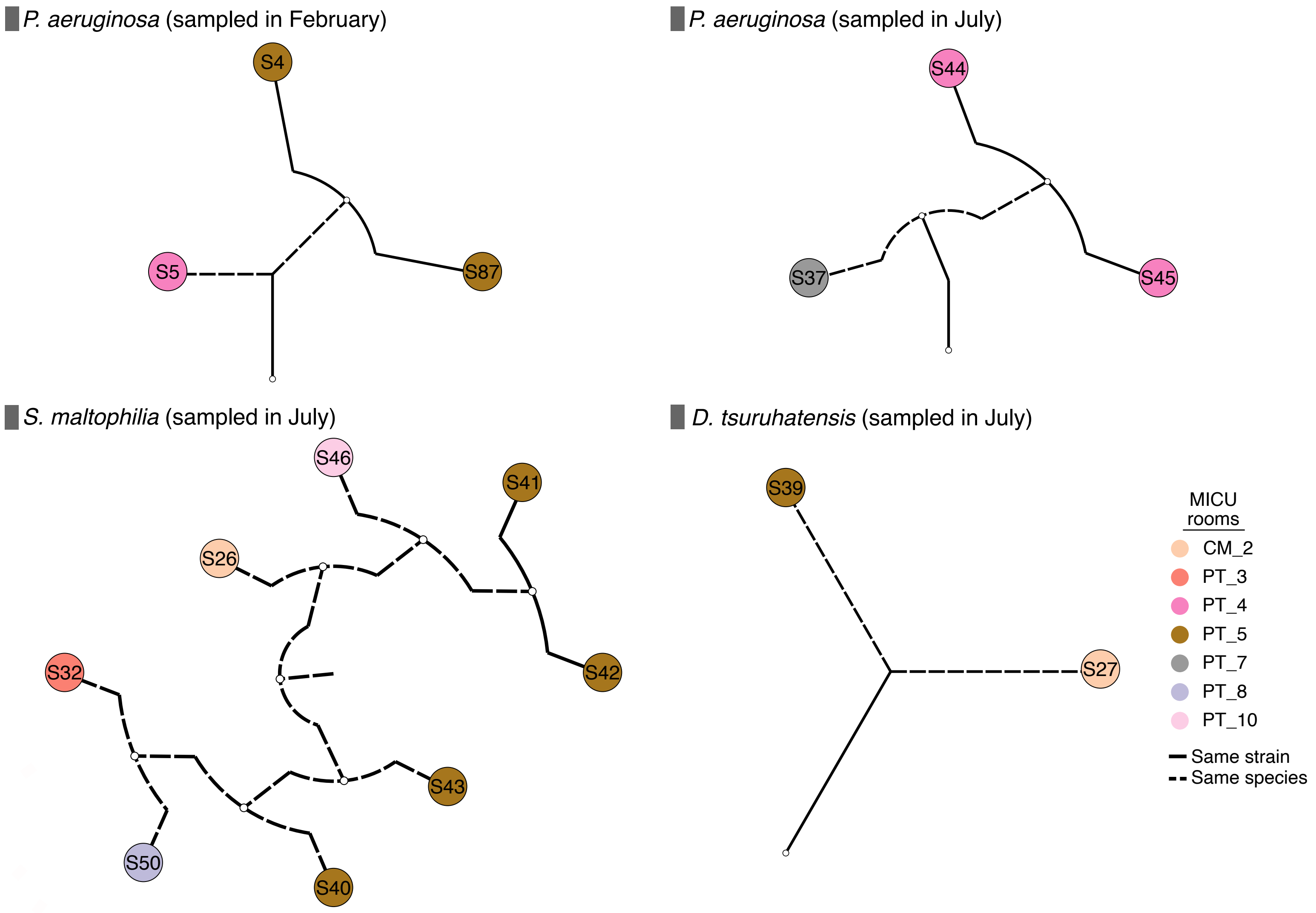


**Fig. S7 Potential dissemination of opportunistic pathogens across multiple MICU rooms.** Patient rooms were deidentified. Each panel shows one species with multiple isolates detected in sink drains of different MICU rooms during the same sampling event.

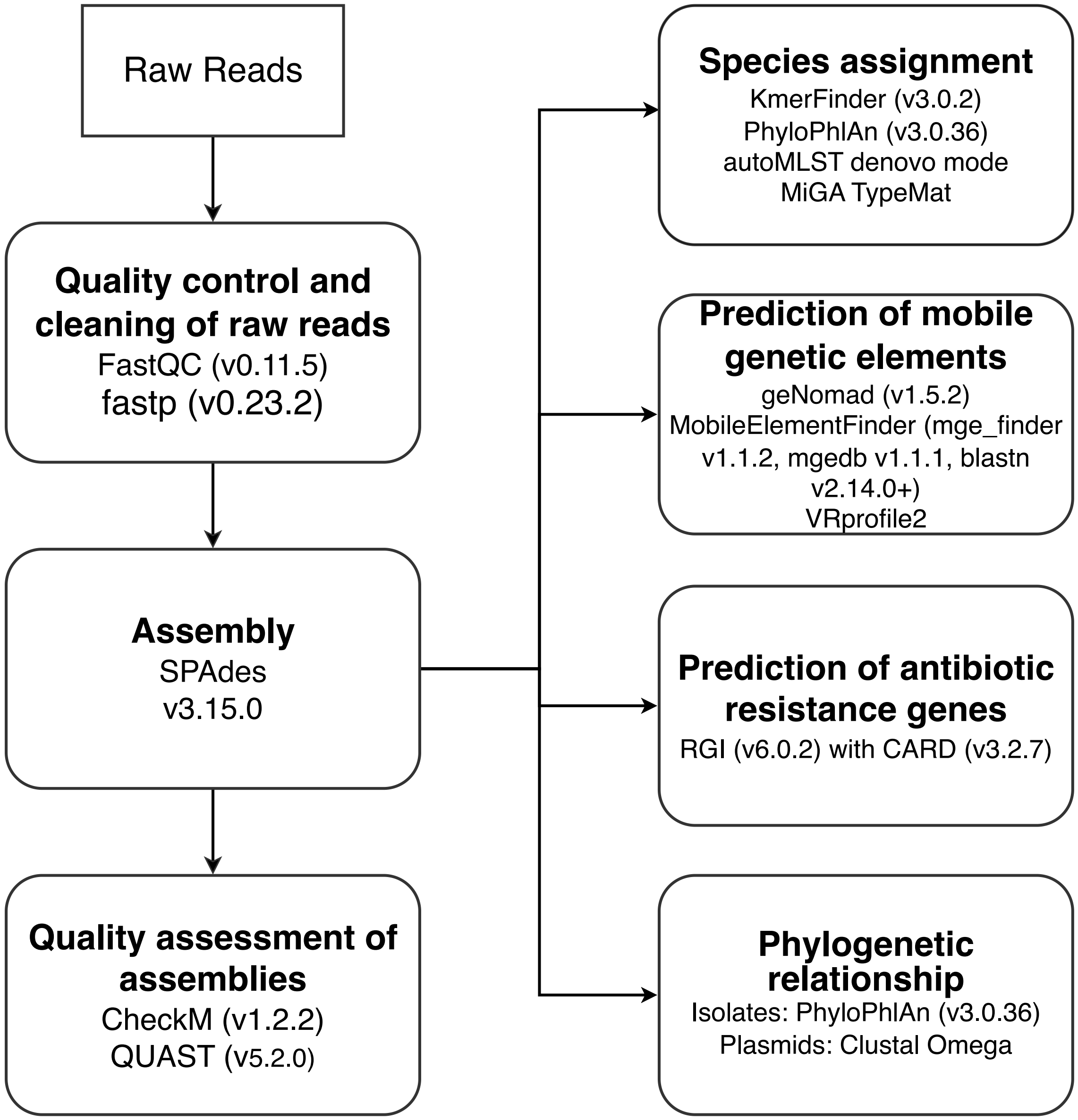


**Fig. S8 Analysis pipeline of whole-genome sequences.**
